# Revisiting TabGraphSyn: training boundaries and record-level conditioning in synthetic clinical data evaluation

**DOI:** 10.64898/2025.12.28.25342851

**Authors:** Zongqian Wu, Haoyuan Cheng, Rahat Khan, Huiping Chen, Jake Y Chen

## Abstract

**Objective:** To investigate how training-set contamination and record-specific conditioning affect utility estimates and membership-inference audits in synthetic clinical data generation, using TabGraphSyn as a case study.

**Materials and Methods:** We combined implementation inspection, metric calibration, controlled changes to released synthetic tables, and train-only experiments across four cohorts containing 683 to 156,468 records. Paired retraining before and after a bundled train-only correction used three shared seeds on TCGA and ACTG 175. We assessed train-on-synthetic, test-on-real (TSTR) utility, two membership-attack families, and the decodability of record content from conditioning vectors.

**Results:** In a contaminated MIMIC-IV output, removing synthetic records seeded by designated holdout records increased the nominal distance-attack excess from −0.0003 to 0.2248 without retraining. After the bundled train-only correction, TCGA TSTR fell by 0.0279 and the per-record GNN arm showed an additional attack signal of 4.0 resolution units relative to no conditioning. Across train-only TCGA arms, condition-to-latent decodability correlated with this signal (Spearman rho = 0.96 across ten experimental arms; exploratory); the corresponding ACTG 175 association was weaker and not statistically significant. A full-data MIA-EPT rerun detected the MIMIC-IV per-record arm after a subsampling-related sensitivity problem was addressed. Among evaluable train-only comparisons, no tested variant combined attack signals within the declared thresholds with a statistically supported utility gain over clean TabSyn.

**Discussion:** Contamination can increase measured utility while weakening the member-versus-non-member contrast on which a privacy audit depends. Record content carried by a conditioning vector provides a separate potential disclosure channel. These mechanisms require distinct controls.

**Conclusion:** Joint utility and privacy evaluation requires verified training boundaries, calibrated attacks and direct examination of conditioning information. Null attack results and non-significant utility differences remain limited by the tested attacks, cohorts and replication.

## 1 Background and significance

Synthetic patient data are proposed as a way to share clinical information without exposing individuals [2–4]. Their value rests on two claims that are usually evaluated together: that models trained on synthetic records perform like models trained on real records, and that the synthetic records do not disclose who was in the training data. The first is commonly measured by training on synthetic data and testing on real data (TSTR) [5], the second by distance-to-closest-record statistics or membership-inference attacks [6–8].

TSTR generalisation and membership inference both depend on correctly identifying records that were excluded from generator training. If designated holdout records enter training, utility estimates can become optimistic and the nominal non-member group no longer provides a valid membership contrast. Leakage is a recognised source of irreproducibility in machine-learning research [9,10], and distance-based privacy metrics alone are insufficient to establish protection against membership inference [11,12]. We examine how these concerns interact within one generator family and how controlled interventions can distinguish them.

Tabular diffusion models learn to generate mixed numerical and categorical records, either directly in feature space [13,14] or through a continuous latent representation [15]. TabGraphSyn adds a graph-derived condition to latent diffusion [1]. Its motivation is that nearby records may carry useful local information about joint feature structure that is not explicitly represented by an unconditional generator. A k-nearest-neighbour graph connects similar records, and a graph isomorphism network (GIN) [16] converts the record and its neighbourhood into a conditioning vector. This is a similarity graph inferred from a single table, not a graph of independently observed relationships between patients. Relational generators designed around links between tables provide the architectural precedent [17,18].

The proposed benefit creates a diagnostic tension. A condition rich enough to preserve local structure may also retain the content of its source record. In addition, a high-fidelity synthetic table can appear useful even when the evaluation records entered generator training. We therefore study TabGraphSyn as a case in which training membership, the conditioning information path and attack sensitivity can be inspected separately. The earlier method report [1] evaluated marginal and pairwise fidelity, schema validity, classification, survival-related covariate recovery, augmentation and privacy proxies. Here, those objectives motivate the diagnosis; their historical numerical results are not treated as independently validated findings. The generator, cohorts, interventions and outcome definitions needed to interpret the present experiments are specified below.

Per-record conditioning raises a further question even when the split is valid. An embedding derived from a real record may retain enough of its content to guide the generator towards that record. Recent work systematically benchmarks membership attacks against tabular synthesis [19] and examines memorisation in tabular diffusion models [20]. We test whether leakage is associated with the information encoded in the condition, using random per-record codes, record latents and cluster-level conditions as controls.

## 2 Objective

We investigate how evaluation contamination and record-specific conditioning interact in synthetic clinical data generation. Four questions organise the study: which records reach each training stage; whether the composition of released synthetic records can suppress a membership contrast; whether record information recoverable from a condition is associated with disclosure after the split is corrected; and whether the tested alternatives offer utility gains over a corrected local TabSyn comparator within declared attack thresholds. The contribution is a mechanism-based diagnosis and evaluation procedure, supported by implementation evidence, interventions and bounded comparisons. This is not a new clinical prediction model or a claim that one generator is uniformly best.

## 3 Materials and methods

### Scope and pipeline definitions

We distinguish three implementation states. The released implementation is the code associated with the December 2025 preprint [1], inspected here for its feature mask and data-loading behaviour. The post-mask contaminated pipeline is the subsequent development version, in which informative per-record conditioning was enabled but the designated holdout remained in training. The train-only pipeline restricts training, conditioning and model selection to training rows. The paired retraining experiments compare the latter two states; they do not reproduce the original preprint’s masked implementation or its exact numerical results. The correspondence between each published result and the code used to produce it remains incompletely established. Appendix A separates implementation findings, claims requiring re-evaluation, and directly checkable reporting corrections.

### Cohorts and splits

We evaluated four single-table clinical cohorts spanning 683 to 156,468 records (Table 1). TCGA is the Pan-Cancer Atlas clinical cohort used for survival modelling [21]. ACTG 175 is the AIDS Clinical Trials Group Study 175 trial dataset [22]; stored experiment files abbreviate it as "AIDS". WBCD is the Wisconsin Breast Cancer (Original) dataset, 683 complete records with nine integer cytology features and a benign/malignant label [23]. MIMIC-IV is a cohort of 156,468 hospital admissions derived from MIMIC-IV [24], with 13 numerical features (age, length of stay, ten laboratory values and days to the follow-up event) and 9 categorical features (including gender, race, insurance, language, marital status, admission type and location, and the in-hospital death flag). Each MIMIC-IV row is one hospital admission, the last for each patient, so no patient contributes more than one row. The in-hospital death flag (1 = died) is the main target. Two further columns record a post-admission outcome, a follow-up death indicator and the number of days to that event; they are removed whenever in-hospital death is predicted from the other columns. Insurance, language and marital status are missing for 2,898, 522 and 8,500 admissions; these gaps are modelled as a separate category during training and written back as missing in the released tables, and the stored training split has no other missing values. MIMIC-IV is a credentialed-access resource, and we report only aggregate statistics.

**Table 1.**
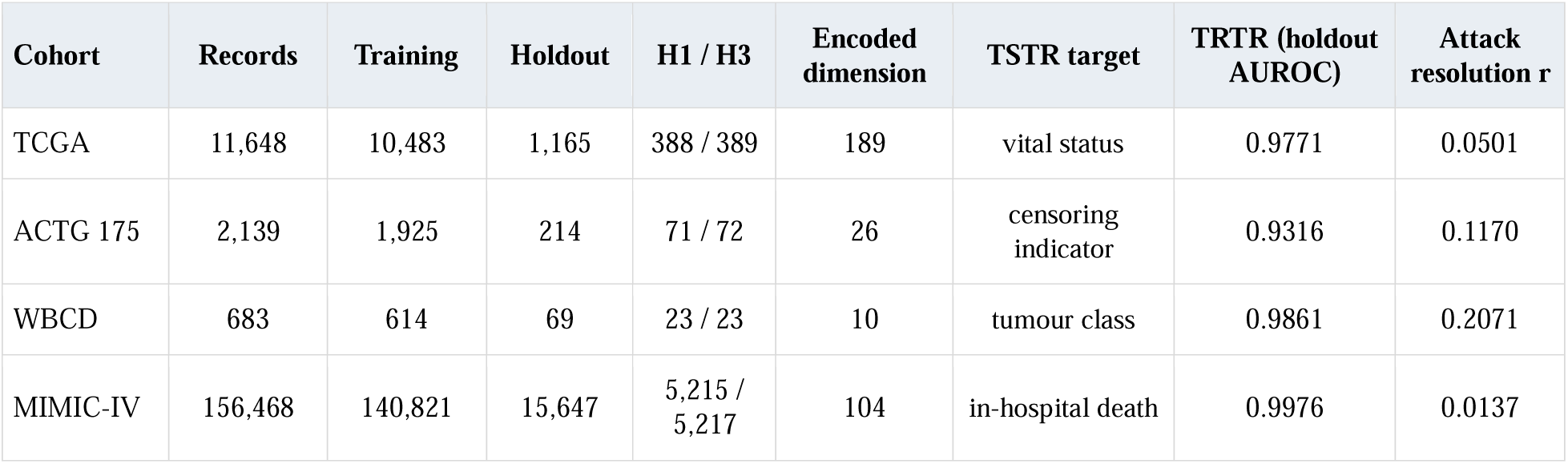
Cohorts and evaluation sizes. H1 and H3 are the first and last thirds of the designated holdout. They serve as non-members and a negative-control subset only when excluded from generator training. Encoded dimension is the width of the distance space. TRTR is the AUROC of a random forest trained on real training rows and tested on the designated holdout. Resolution is the declared approximate null half-width for attack excess (Methods). Six additional MIMIC-IV targets address saturation of in-hospital death prediction (Figure 4).

| **Cohort** | **Records** | **Training** | **Holdout** | **H1 / H3** | **Encoded dimension** | **TSTR target** | **TRTR (holdout AUROC)** | **Attack resolution r** |
|---|---|---|---|---|---|---|---|---|
| TCGA | 11,648 | 10,483 | 1,165 | 388 / 389 | 189 | vital status | 0.9771 | 0.0501 |
| ACTG 175 | 2,139 | 1,925 | 214 | 71 / 72 | 26 | censoring indicator | 0.9316 | 0.1170 |
| WBCD | 683 | 614 | 69 | 23 / 23 | 10 | tumour class | 0.9861 | 0.2071 |
| MIMIC-IV | 156,468 | 140,821 | 15,647 | 5,215 / 5,217 | 104 | in-hospital death | 0.9976 | 0.0137 |

The MIMIC-IV cohort was built from version 3.1 of the hosp module. Admissions were joined to patient records, and for each patient only the admission with the latest discharge time was kept. Ten routine laboratory values (glucose, creatinine, urea nitrogen, white blood cell count, haemoglobin, platelet count, sodium, potassium, bicarbonate and haematocrit) were averaged over all results linked to that admission. Admissions with a non-positive recorded length of stay or lacking at least one of the ten laboratory values were excluded; the laboratory criterion removed 66,860 of 223,328 last admissions, leaving 156,468 admissions from as many patients. Age is the MIMIC-IV anchor age. The follow-up death indicator marks death in hospital or within 365 days of discharge, and the days to that event are capped at 365, the value also given to censored patients.

Each cohort has a fixed 90/10 split into training and designated holdout rows, stored with the code. The split is defined by a stored index permutation: its first n_train entries are training rows and its last n_holdout entries the designated holdout. The repository’s preprocessing script generates this permutation by shuffling with seed 1234 and incrementing the seed until every categorical level occurs among the training rows; we used the stored files without regenerating them. The term designated holdout refers to these evaluation indices; the records are genuinely unseen by the generator only in the train-only arms. For the distance attack, the designated holdout is divided in stored order into contiguous subsets H1, H2 and H3 (D4, Appendix B). H2 was reserved as the reference set of a density-ratio attack that was discarded after failing its positive control (D4); the distance attack does not use it. Distances are Euclidean after one-hot encoding of categorical columns and standard scaling (see Membership attacks). Storage type determines which columns are treated as categorical, so integer-coded categories are treated as numeric. The resulting dimensions are 189 for TCGA, 26 for ACTG 175, 10 for WBCD and 104 for MIMIC-IV. The reported splits and attacks operate at the row level; the MIMIC-IV rows are admissions.

### Generator design and information flow

TabGraphSyn consists of a tabular variational autoencoder (VAE), a similarity-graph encoder and a conditional latent diffusion model [1,15,16,25]. A record x_i contains numerical and categorical attributes.

The VAE encodes it into a continuous vector z_i and decodes generated vectors back into the original table schema. Its reconstruction and latent-regularisation objectives allow diffusion to operate in a continuous representation while the decoder handles mixed data types. Numerical inputs use quantile normalisation. The graph encoder and diffusion model are separate learned stages; their inputs and training membership must therefore be checked separately (Figure 1).

**Figure 1.**
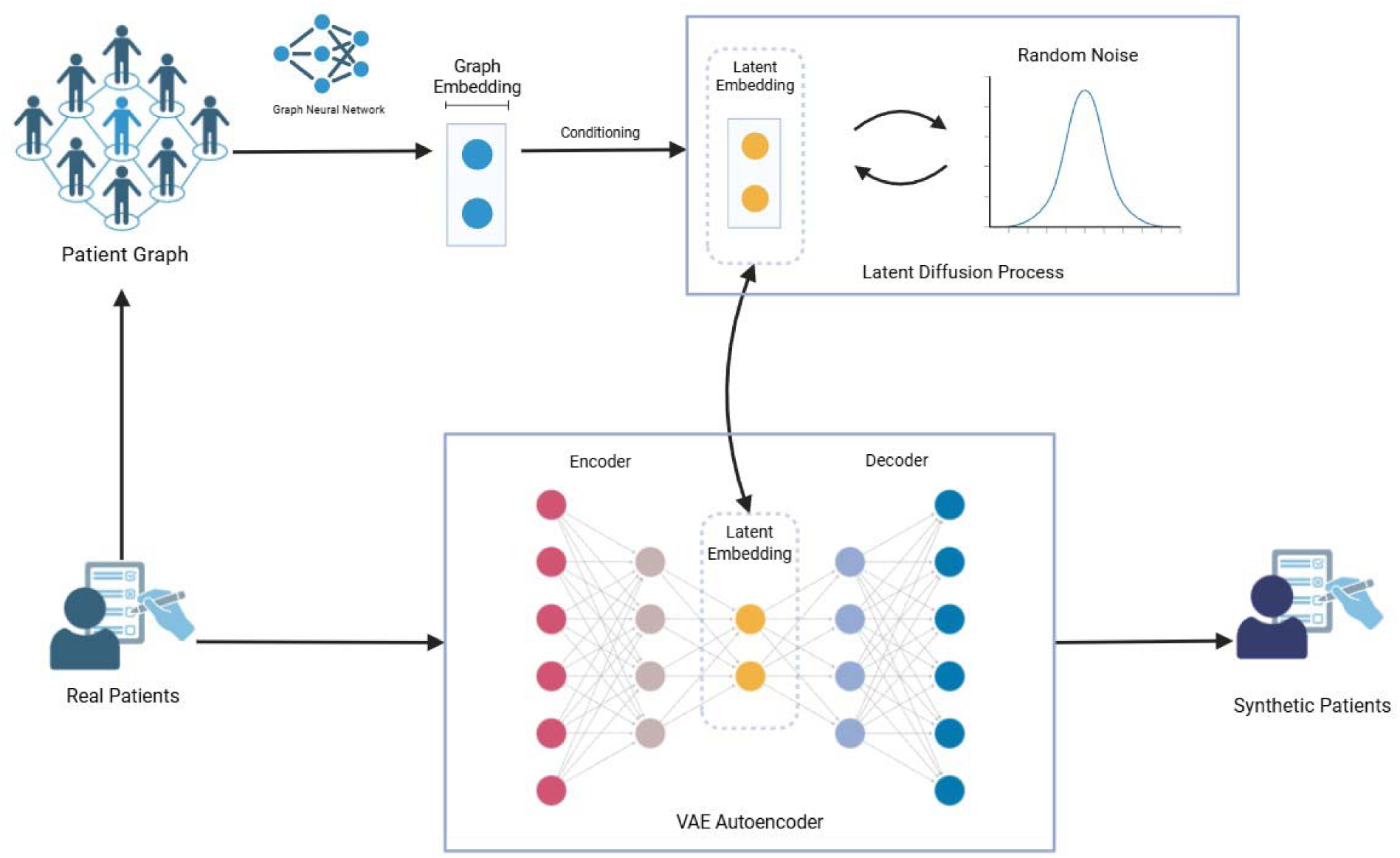
TabGraphSyn’s intended information flow, shown here in full [1]. A VAE produces record latents; a similarity-graph encoder produces conditions; conditional diffusion and VAE decoding produce synthetic records. The diagram describes the architecture rather than proving which information path was active in a particular run. The released-code mask removed record-specific graph inputs. In the post-mask contaminated pipeline, designated holdout rows entered latent training, graph construction, conditioning and model selection. The train-only correction restricts those stages to training rows.

Graph construction and encoding. Each record is a node, and nearest-neighbour edges represent feature similarity within the available record pool. With informative inputs enabled, the GIN aggregates the record’s own features and neighbouring features. It is trained to predict that record’s VAE latent z_i, and its hidden vector h_i is cached as the condition. Thus, neighbourhood information and the source record’s own information can both reach h_i. The presence of graph edges does not by itself establish that any subsequent gain comes from relationships among records rather than from encoding the record itself.

Conditional diffusion and sampling. Gaussian noise is added to record latents during training, and the denoising network learns to recover latent structure conditional on h_i under the diffusion formulation [25]. Generation starts from noise, applies 100 denoising steps with a selected condition, and decodes the resulting latent to a synthetic row. The condition therefore enters the generative process in addition to the random noise. The output is a new draw from a learned conditional distribution; it is not guaranteed to be independent of the real record that supplied its condition.

Condition selection is part of the model. For a cached pool of N conditions, the sampling distribution can be written conceptually as p_syn(x) = (1/N) sum_i p_theta(x | h_i), with decoding included in p_theta. The implementation selects cached conditions without replacement when the requested output size does not exceed the pool. A release with N rows consequently uses each condition once. In the contaminated post-mask pipeline, N includes designated holdout rows; in the train-only pipeline, it includes training rows only. The cluster-conditioned variants replace these empirical point conditions with draws from a fitted mixture. This distinction is central to the output-removal and conditioning-content experiments.

The TabGraphSyn backbone used k = 5 neighbours, GNN hidden dimension 128, a multilayer-perceptron denoiser, quantile normalisation, 100 denoising steps, and maximum training lengths of 4,000 VAE, 1,000 GNN and 10,000 diffusion epochs with the repository’s early stopping. The initial preprint stated k = 3, whereas the inspected implementation uses k = 5. Baselines follow their own model specifications. We report this discrepancy for reproducibility; we did not conduct a general sensitivity analysis over these settings. Settings not listed here are the defaults of the inspected code version; experiment configurations are documented in the declarations in Supplementary Data S1.

### Conditioning information and training boundaries

Feature masking. In the source multi-table generator, a table’s own features are hidden while its embedding is computed [17]. In the inspected released single-table code, the only table remained in the masked list. Node features and the inputs to graph construction were therefore constant. Under the implemented fixed-degree aggregation, this removes patient-specific variation from the embeddings. The relevant released files are unchanged from the first commit on 29 August 2025. The mask was removed on the development branch on 16 February 2026 (commit 672f797). The quantitative experiments analysed here use post-mask outputs or train-only variants. The earlier masked state is assessed through code and working records rather than a new reproduction run.

Per-record conditioning. After masking is removed, the GNN receives the record’s own content and learns to predict its VAE latent. Sampling from its cached embedding consequently provides information specific to a real record. We test the disclosure implications of this design rather than treating the presence of a unique condition alone as evidence of leakage.

Training-set contamination. The inspected diffusion loader read the all-row latent file (latents.npy) rather than training-row latents (train_z.npy). The graph, GNN and conditioning pool also included all rows, and VAE model selection used an internal validation split from the full table. The local TabSyn adaptation contained the same all-row diffusion-loading path. Inspection of upstream TabSyn (amazon-science/tabsyn, commit cb5ac0f) showed training-only VAE and diffusion inputs [26]; the local loading problem should not be attributed to the published TabSyn method. Saved-run evidence and the limits of inference from the initial preprint’s output files are reported separately in Results.

The train-only correction restricts the latent inputs, graph, GNN and conditioning pool to training rows. VAE model selection uses an internal 10% split of those training rows. The correction comprises 20 hunks across four files, including run isolation and alignment checks. Stored latent means were compared with all-row and training-row means to check saved-run provenance. The paired experiments evaluate this pipeline-level correction; they do not isolate the contribution of each changed component.

### Evidence sources

Evidence comes from code history, dated working records, saved outputs and the controlled experiments reported here. The historical output collection contains 233 configurations from 11 mitigation families, including sampling-time and training-time perturbations, embedding mixup, neighbour-count changes and augmentation. It provides context and candidate outputs for diagnosis, not 233 independent confirmations of a single hypothesis. Values taken from working records without recomputation are marked in Table 2. The initial preprint’s exact results were not regenerated.

**Table 2.** Diagnostic checkpoints and evidence boundaries. The marked input-vector count is from a February 2026 working record and was not recomputed. Historical novelty and fidelity comparisons are described in Results with their concurrent changes. Details needed to reconcile the initial preprint are retained in Appendix A.

| Checkpoint | Implementation state | Evidence | Diagnostic implication | Boundary |
|---|---|---|---|---|
| Condition varies across records | Released masked code | Constant graph inputs; historical WBCD log: 1 distinct vector before repair, 449 after* | Patient-specific graph information is absent in the inspected masked implementation | No new masked-code rerun; original result provenance incomplete |
| Condition carries record content | Post-mask design | Own features enter the GNN; latent-regression objective; decodability and identity-content controls | A record-specific information path can persist after the split is corrected | Association and controls concern the tested model family |
| Evaluation rows remain unseen | Contaminated post-mask pipeline | Loader inspection, saved latent means, condition-to-row mapping and paired retraining | Nominal holdout exposure compromises both TSTR and membership contrasts | Correction bundles several training-boundary and run-control changes |
| Attack responds to known disclosure | Legacy suite and later attack variants | Copy calibration, non-member controls and MIA-EPT sampling-overlap checks | A favourable metric needs a valid membership definition and representative controls | Copy detection alone does not establish sensitivity to every release |
| Comparator reads the intended rows | Local TabSyn adaptation | Local all-row loader differs from inspected upstream training-only code | Baseline adaptation and membership boundary require independent checks | Output size alone does not establish exposure of historical baseline files |

Table 2 connects each diagnostic checkpoint to the evidence and the inference it permits. Implementation, sampling and evaluation issues are distinguished rather than assumed to form a completely identified historical causal sequence.

### Experimental arms

Experimental arms were trained in separate code copies to preserve the source implementation. Table 3 defines their training and sampling inputs.

**Table 3.** Experimental arms. All arms except the contaminated post-mask arm use training rows only. Standard seeds are 42, 123 and 456. MIMIC-IV clipping was added at gradient norm 1.0 for the matched cluster-level and no-conditioning reruns after four of ten initial trainings diverged.

| Arm | Conditioning during training | Conditioning during sampling | Cohorts | Seeds |
|---|---|---|---|---|
| Contaminated post-mask | GNN embedding of the record; holdout rows in every training step | Embedding of a real record (all rows) | TCGA, ACTG 175 (retrained); MIMIC-IV (2 existing outputs) | 3; — |
| No conditioning | None | None | All | 3 (MIMIC-IV 2 + 3 with clipping) |
| GNN embedding (per-record condition) | GNN embedding of the record | Embedding of one training record | All | 3 (MIMIC-IV 1) |
| Record's own latent | VAE latent of the record | Latent of one training record | TCGA, ACTG 175 | 3 |
| Random per-record code | Random vector fixed per record | Code of one training record | TCGA, ACTG 175 | 3 |
| Cluster-level hint p(h) | Draw from the record's mixture component (rule-based K) | Draw from the mixture | All (MIMIC-IV with clipping) | 3 |
| Cluster-level hint, K = 1–2,048 | Draw from the record's K-means component | Draw from the mixture | TCGA (K up to 2,048), ACTG 175 (up to 512) | 3 (K = 1: 1) |
| Latent-based partition | As p(h); VAE-latent groups, GNN-space conditions | Draw from the mixture | All | 3 (MIMIC-IV 1) |
| Graph used only during training | None; batches mixed with latent-space neighbours (p = 0.5 or 1.0) | None | TCGA | 3 per setting |
| Clean TabSyn | None (TabSyn adaptation, two declared fixes) | None | All | 3 (MIMIC-IV 1 converged) |
| Published baselines | TabDiff, TabDDPM, CTGAN, CTAB-GAN+ on training rows | — | MIMIC-IV | 1 |
| Verbatim copy (positive control) | Training rows submitted as the synthetic table | — | All | — |

Contaminated post-mask arm: informative GNN conditions with designated holdout rows included in training and sampling. TCGA and ACTG 175 were retrained with the same seeds and settings as the train-only GNN arm. MIMIC-IV comparisons use two existing contaminated outputs.

Per-record conditions in the train-only pipeline: the record’s GNN embedding, a representation of its own VAE latent, or a random vector drawn once per record. The record-latent control uses standardised principal components of the VAE latent (PCA-64); the random-code control uses Gaussian codes. Both are scaled to match the GNN conditions. The latter distinguishes records without directly encoding their features; comparing these arms tests recoverable content as well as uniqueness.

Cluster-level conditions p(h): each record is assigned to a component c(i). During training, its condition is freshly sampled from that component’s Gaussian distribution with mean mu_c and covariance Sigma_c, while the target latent remains z_i. During generation, a component is drawn according to its fitted mixture weight and a condition is sampled from that component, rather than selected from an individual record’s cached vector. The rule-based arm partitions GNN embeddings with a Gaussian mixture model (GMM); the granularity experiments use K-means partitions with requested K = 128, 512 or 2,048 where specified in Table 3, alongside existing K = 1 and rule-based endpoints.

The rule-based GMM selects K_lik from {1, 2, 4, 8, 16, 32} by likelihood on an internal 10% split of training conditions, caps K at floor(n_train/100), and merges components smaller than 100 records. It uses full covariance, regularisation 10^-4 and random state 42; the effective component count after merging is reported in Figure 6. The K-means ladder uses three initialisations, random state 42 and component covariance regularisation 10^-4 I; singleton components are merged into the nearest centroid. These protocols vary both granularity and partitioning procedure, so the ladder is not a fully isolated intervention on K alone.

Latent-based partition control. A separate GMM assigns records to groups using standardised VAE latents after PCA-64 rather than GNN embeddings. The Gaussian condition means and covariances are still calculated in GNN-embedding space. This control changes which records are grouped together; it does not remove the GNN from the conditioning representation. We call it the latent-partition control throughout.

Graph information used only during training: in each clean TabSyn training batch, a record’s latent z_i is replaced with probability p by λz_i + (1 − λ)z_j, where j is drawn uniformly from the five nearest training-latent neighbours of i (Euclidean distance on normalised latents) and λ ∼ Uniform(λ_min, 1). The two declared settings were p = 0.5 with λ_min = 0.5, and p = 1.0 with λ_min = 0 (D12). Sampling is unconditional.

No conditioning: the same backbone without a conditioning vector.

Clean TabSyn: the local TabSyn adaptation with two declared fixes, restricting diffusion inputs to training-row latents and passing the parsed seed to training. This is a corrected local comparator, not a claim to have reproduced every setting of the upstream benchmark.

Additional baselines: TabDiff [14], TabDDPM [13], CTGAN [27] and CTAB-GAN+ [28], trained on MIMIC-IV training rows only (D13).

Train-only arms used seeds 42, 123 and 456 unless stated otherwise and generated the training-set number of records. The contaminated post-mask arm used the all-row conditioning pool and full-table sample size. Replicate counts and convergence exclusions are stated in Table 3 and Results.

Replication and shared representations. In the declared TCGA, ACTG 175 and WBCD train-only experiments, each cohort’s VAE was trained once and shared across the conditioning arms; GNN and diffusion seeds provide the stated repetitions. The contaminated paired comparator similarly shares its seed-42 VAE across its later seeds. The granularity and identity-content experiments reuse the previously trained representations. MIMIC-IV repeats reuse the seed-42 graph representation where applicable, so additional diffusion seeds are not independent end-to-end model replications. Seed-based uncertainty therefore describes variation conditional on these representations and the fixed data split.

### Measurements

Evaluation setting. A synthetic table is the released object. Utility asks whether a classifier trained on that table transfers to real evaluation records. Membership inference asks whether a candidate real record participated in generator training, using its relationship to the released table. The distance attack requires candidate record attributes and the synthetic table; the MIA-EPT experiment additionally uses labelled attack-training examples. Source indices, checkpoints and latent means are available to the investigator for mechanism diagnosis, not assumed to be available to an ordinary recipient. The results are empirical disclosure tests under these access conditions, not a formal privacy guarantee or an exhaustive assessment of all attacks.

Utility. A random forest with 100 trees and seed 42 was trained on synthetic data and scored by AUROC on the fixed designated holdout (TSTR) [5]. The same classifier trained on real training rows gives TRTR. TRTR is a model-specific empirical reference, not a theoretical upper bound. Targets are vital status for TCGA, the censoring indicator for ACTG 175, tumour class for WBCD, and in-hospital death for MIMIC-IV. Because the last is saturated (TRTR 0.998), D11 added six non-saturated tasks: in-hospital death after removal of post-admission outcome columns, and gender, insurance, marital status, admission type and admission location, each predicted from the remaining columns. For binary targets AUROC uses the predicted probability of the positive class; for multiclass targets it is the one-vs-rest macro average over the classes present in the real training rows (scikit-learn 1.4.2), with probabilities from the synthetic-trained classifier aligned to those classes. Records with a missing target are excluded from training and testing, and missing categorical predictors are encoded as all-zero indicators. These tasks assess information preservation rather than prospective clinical validity. For historical comparison, we also applied the earlier evaluation protocol, which re-splits the full table after generation. That protocol does not provide an independent test for an all-row-trained generator.

Fidelity. We report the mean two-sample Kolmogorov–Smirnov statistic over numerical columns and the root-mean-square error between real and synthetic pairwise Pearson correlations, both computed against the holdout rows, and the median distance from each synthetic record to its closest training record. Lower KS and correlation error indicate closer agreement with the reference distribution. These fidelity measures do not establish clinical plausibility or privacy. Schema validity means preserving variable types and allowed formats; it is distinct from preserving treatment effects, survival processes or valid combinations of clinical attributes.

Membership attacks. The primary score is the negative Euclidean distance from each real record to its nearest synthetic record. For each contrast, categorical columns are one-hot encoded and all columns are standardised with one scaler fitted on the union of the two candidate sets and the synthetic table; missing values become zero after encoding. The main AUC compares training rows with H1; the control AUC compares H3 with H1 (D4). In train-only arms, H1 and H3 are both non-members, providing an empirical negative control. In contaminated arms they were exposed to training, so the same calculations describe nominal split contrasts rather than a valid membership test. Attack excess E is the main AUC minus the control AUC. The declared resolution is r = 1.96 × √[v(n_train, n_H1) + v(n_H3, n_H1)], where v(a, b) = (a + b + 1)/(12ab) is the Hanley–McNeil variance of an AUC of 0.5 between groups of sizes a and b with continuous, untied scores [29]: 0.0501 for TCGA, 0.1170 for ACTG 175, 0.2071 for WBCD and 0.0137 for MIMIC-IV. The calculation treats the main and control AUCs as independent although both use H1, assumes no tied scores, and ignores the uncertainty of the no-conditioning excess subtracted below. The shared H1 would be expected to correlate the two AUCs positively, which makes r conservative for E alone, whereas the estimated reference adds variance; r is therefore a null scale for E, not a confidence interval for D or for differences between arms. Exact zero distances (WBCD) depart further from these assumptions. Supplementary Table S1 gives the main AUC, control AUC, E and D of every analysed run. We additionally report D = (E_arm - mean E_no-conditioning) / r. Values within +/-1 meet the study’s operational threshold for no resolved additional signal relative to the backbone; they do not establish absence of absolute leakage. This scale is an approximate calibration, not a formal privacy bound. WBCD is not sufficiently informative for this comparison: 30% of holdout rows duplicate training rows, and the copy control reaches only 0.72 units of D (raw E = 0.2817; backbone E = 0.1330).

Reading the attack scale. AUROC is the area under the receiver operating characteristic curve: 0.5 corresponds to chance ranking and values above 0.5 indicate better member ranking under the specified score. E removes the empirical control contrast, while D additionally subtracts the no-conditioning backbone and divides by the declared resolution r. For example, TCGA D = 4 means that the arm’s excess is approximately 4 x 0.0501 = 0.2004 AUC units above the backbone excess; it is not a fourfold probability of identifying a person. Likewise, D near zero means similar measured signal to that backbone, not necessarily an absence of membership information.

The second attack family, membership inference via error prediction for tabular data (MIA-EPT) [30], uses per-column attribute predictors trained on synthetic data and an attack classifier on their error profiles. We used the released feature construction with the recorded adaptations in D5: direct labelled attack training without shadow models, a gradient-boosting classifier, runtime fixes, and a 20,000-row cap on MIMIC-IV predictor training. High-cardinality text classification and MIMIC-IV missing-value handling were added after earlier cohorts had produced partial results, before results existed for the affected cohorts; earlier cohorts were not rerun. These are adaptations rather than an exact reproduction of the published attack. Ground-truth membership is available to the evaluator; practical attacker access is not established by this experiment. The holdout is split in stored order into halves A and B, and members M are sampled without replacement from the training rows to size |A| (seed 42). Per-column attribute predictors are fitted on the synthetic table only; the challenge records in M, A and B are used only to compute error profiles. The attack classifier (histogram gradient boosting with library defaults and seed 42; no hyperparameters were tuned) is evaluated by stratified five-fold cross-fitting: each challenge record is scored by a classifier trained on the other four folds, and the AUC is computed from the pooled out-of-fold scores. The main comparison is M versus B and the control is A versus B, so B supplies the negatives in both; the MIA-EPT resolution applies the formula above with both terms equal to v(|B|, |B|). The MIMIC-IV cap and subsequent full-data rerun are examined explicitly in Results.

Positive controls. We submitted the training rows as a verbatim-copy synthetic table for each cohort and attack. Failure to detect this control limits interpretation of a null result. Successful detection checks sensitivity to that control but does not ensure equal sensitivity to other release mechanisms or sampling schemes.

Condition decodability. Five-fold out-of-fold ridge regression predicts each training record’s VAE latent from its training condition; we report the mean per-dimension R-squared (D11). The ridge penalty is chosen from {0.1, 1, 10, 100, 1000} by efficient leave-one-out validation within each training fold; conditions and latent dimensions are standardised once over all training records before the folds are formed (fold seed 0), and no PCA is applied. For cluster-level conditions the predictor is the fitted mean of the record’s component. The no-conditioning arm is assigned R-squared = 0 and D = 0 by definition.

These are diagnostics of already-fitted representations, not tests of generalisation to new patients. This measures one form of linear record-content recoverability; it is not a complete information measure and does not capture every property of the component distribution.

### Calibration of the project’s metrics

The project’s evaluation suite reported a membership-inference AUC, a distance-to-closest-record statistic, a detection score, TSTR, and the NewRowSynthesis metric of the SDMetrics library, version 0.27.0 [31, 32]. We applied the first two to generators with known ground truth: an exact or near-duplicate copy of the training data, the copy with added Gaussian noise, an independent sample from the same distribution, and an over-dispersed sample.

Historical perturbation controls. The saved output-removal comparisons include unperturbed conditions, sampling-time perturbations and training-time perturbations. The latter two add noise to cached conditions along directions estimated by local principal-component analysis of neighbouring conditions. Sampling-time perturbation changes conditions only during generation; training-time perturbation redraws perturbed conditions during model fitting while retaining the source record’s target latent. These settings probe sensitivity to condition variation. Their presence in the output collection does not make them validated privacy protections, and the present intervention does not separately estimate the effect of every perturbation setting.

Output removal intervention. When the contaminated pipeline generates one output per full-table row, every designated holdout row also seeds an output. We mapped synthetic records to their conditioning source and removed outputs seeded by designated holdout records, then repeated the distance contrast (D7). Unconditional outputs underwent equal-sized random removal. The trained models were unchanged and still contained holdout information. This intervention isolates an output-composition effect; it is not a train-only retraining experiment or a formal lower bound on leakage of a differently trained model.

### Declarations and statistical analysis

For the 12 diagnostic experiments D1-D12, written declarations recorded arms, measurements, predictions and decision rules (Appendix B), and their SHA-256 hashes are listed in Supplementary Data S1. Ten were frozen before launch; two were saved 33 and 77 seconds after launch, before reported outcomes were available. The latter launch failed out of memory and was rerun hours later. D13 separately records the MIMIC-IV baseline runs. These are dated internal declarations rather than an assertion of public prospective registration. Training runs whose best diffusion loss occurred before epoch 200, or whose early-stopping loss exceeded 0.9, were excluded as diverged. Four of ten initial train-only MIMIC-IV trainings diverged; matched cluster-level and no-conditioning arms were rerun with clipping and all six converged. Two of three clean TabSyn MIMIC-IV seeds also diverged.

Welch two-sided t-tests compare independent arms over seeds, and paired t-tests compare contaminated and train-only runs sharing seeds. Seed bootstrap intervals use 10,000 resamples. With usually three seeds, uncertainty estimates are coarse and power is limited; non-significant differences are not equivalence tests. Seed variation conditional on one data split does not quantify uncertainty over populations or alternative splits. Prespecified decision thresholds are distinguished from descriptive comparisons, and the reported comparison p-values should not be read as a universal ranking across all arms. Analyses used Python 3.10, PyTorch 2.0.1 and NVIDIA A100 GPUs.

Decision criteria and comparators. D1-D13 are identifiers for dated experiment declarations, not references to a previous article; Appendix B summarises their scope and outcomes. The initial cluster-condition decision used MIMIC-IV additional attack excess no greater than its declared resolution, with KS and correlation error no more than 0.005 above matched no-conditioning controls. Its TCGA utility criterion required a positive TSTR difference from no conditioning with a one-sided Welch p < 0.05 and additional attack excess within resolution. The later granularity comparisons were exploratory and could not reverse that decision. Graph-only mixup used a separate gain threshold of 0.010 and p < 0.025. Comparisons against clean TabSyn answer a different question and use the reported two-sided Welch tests. We retain these distinctions rather than applying a single post hoc success rule to all experiments.

## 4 Results

### Checking whether conditioning contains record information

Inspection of the released implementation identified constant inputs to graph construction and aggregation. A historical WBCD log recorded one distinct input vector among 683 records before masking was removed and 449 afterwards (Table 2). This supports the conclusion that the inspected masked implementation could not use patient-specific similarity information. It does not independently establish which executable state produced every result in the initial preprint. Historical NewRowSynthesis values, the share of outputs matching no real record, were 1.00 for ACTG 175 and TCGA and 0.41 for WBCD before the repair. These values describe record matching, not privacy guarantees.

### Calibrating novelty and membership measurements

A historical comparison after removal of the mask reported lower mean KS statistics: 0.023 to 0.011 for ACTG 175, 0.009 to 0.003 for TCGA, and 0.032 to 0.010 for WBCD. Novel-record shares fell from 1.00 to 0.28, 1.00 to 0.06, and 0.41 to 0.09, respectively. These working-record values were not recomputed. The same release changed latent scaling and VAE training, preventing attribution of the changes to masking alone. The combination of improved fidelity and reduced novelty motivated direct tests of record-specific conditioning and evaluation sensitivity.

Calibration revealed that several evaluation scores could not answer the intended privacy question (Table 4). The legacy membership metric randomly divided real records into nominal members and non-members without reference to generator training. On simulated data it scored a near-copy at 0.466 and an independent sample at 0.481; using actual membership labels gave 1.000 and 0.516. The legacy distance statistic scored a copy at 0.245 and an independent sample at 0.498 in the calibration, showing that its numerical scale required empirical interpretation. The detection score measures real-versus-synthetic distinguishability and can reward copying. These findings explain why favourable fidelity and detection scores cannot substitute for a calibrated membership audit.

**Table 4.** Behaviour of the project’s evaluation metrics on generators with known ground truth. Membership-inference values are from simulated tables (10 features, 2,000 records); distance-statistic values are means over three simulated cohorts matched to TCGA, ACTG 175 and WBCD. Entries without numbers follow from the metric’s definition.

| **Metric in the evaluation suite** | **How it was computed** | **Copy of the training data** | **Independent sample** | **Consequence** |
|---|---|---|---|---|
| Membership-inference AUC | The real table was split at random into halves labelled members and non-members | 0.466 (near-duplicate copy) | 0.481 | Random nominal labels fail to represent actual membership; the calibrated contrast remains near chance. |
| Distance-to-closest-record share | Share of holdout records closer to the training set than a synthetic record is; 0.5 taken as ideal | 0.245 (exact copy); 0.382 with 0.5 SD noise | 0.498 | The observed scale depends on the metric definition and reference sets; values above 0.5 are not privacy guarantees. |
| Detection score | One minus the advantage of a classifier separating real from synthetic records | Best possible, by construction | Best possible | Measures indistinguishability; a copy is rewarded |
| TSTR | The full table was re-split after generation and the test fold used for scoring | — | — | In the contaminated post-mask pipeline every test record had been used to train the generator |
| NewRowSynthesis | Share of synthetic records matching no real record | 0, by construction | Close to 1 | Detects exact matching under its definition; novelty alone does not establish privacy. |

### Tracing nominal holdout exposure through the pipeline

Saved latent means matched all-row training in 94 of 95 ACTG 175 runs, 99 of 100 TCGA runs, 86 of 87 WBCD runs and all 44 MIMIC-IV runs from the inspected development repository. Remaining cases were indeterminate under the tolerance; none matched training-only means. Saved train-only runs matched the intended training subset (Table 5). Available local TabSyn runs on WBCD and MIMIC-IV also matched all-row training. By contrast, the initial preprint’s supplied baseline outputs have full-table sizes of 11,648, 2,139 and 683 rows, but sample counts alone do not establish their training membership.

**Table 5.** Rows actually used in training, by saved run. Each run’s stored latent mean was compared with the mean over all rows and the mean over training rows. Counts are runs matching each set; the remaining runs were indeterminate under a strict tolerance and none matched the other set. MIMIC-IV runs were assigned by a nearest-candidate re-test.

| **Saved-run source** | **TCGA** | **ACTG 175** | **WBCD** | **MIMIC-IV** |
|---|---|---|---|---|
| Development repository | all rows, 99 of 100 | all rows, 94 of 95 | all rows, 86 of 87 | all rows, 44 of 44 |
| Train-only pipeline | training rows, 28 of 28 | training rows, 28 of 28 | training rows, 26 of 26 | training rows, 16 of 16 |
| Available local TabSyn runs | no saved run | no saved run | all rows, 1 of 1 | all rows, 1 of 1 |

The conditioning-source probe (D6) found 156,468, 11,648, 2,139 and 683 distinct conditions in the MIMIC-IV, TCGA, ACTG 175 and WBCD arrays, matching the full-table row counts. In these outputs each designated holdout row also supplied a condition. With source-to-output correspondence available to the evaluator, the output seeded by a record was its nearest synthetic neighbour for 80% of training and 80% of holdout records on MIMIC-IV, and 74% and 68% on ACTG 175, compared with 1/n under random correspondence. This is a source-linked proximity result under an informed evaluation, not proof that every patient was reconstructed exactly or that an external attacker has the mapping.

### Testing masking by removing holdout seeded outputs

Earlier attacks had used the designated train/holdout labels on contaminated outputs (D4-D5). The distance attack did not exceed its cohort-specific resolution for any of 15 configurations. MIA-EPT exceeded resolution for one ACTG 175 configuration without perturbation (0.130 versus 0.110) and none on the other cohorts. Copy controls were detected. Because the generator had seen both nominal membership groups, these near-null contrasts could not establish privacy of the contaminated outputs.

Removing holdout-seeded outputs changed that contrast without retraining (Figure 2). Across 13 converged MIMIC-IV configurations, distance-attack excess changed from −0.0025 to +0.0017 before removal to 0.047-0.225 afterwards. For the unperturbed configuration it increased from −0.0003 to 0.2248. The unconditional control remained near 0.003 after equal-sized random removal. On ACTG 175, excess increased in all 13 configurations, from at most 0.098 to as much as 0.332, while five unconditional outputs remained within resolution. Among MIMIC-IV configurations meeting the declared fidelity margin, the smallest post-removal excess was 0.101, or 7.4 resolution units. The intervention supports masking by the presence of source-linked outputs for both nominal groups. It does not remove prior training exposure of the holdout.

**Figure 2.**
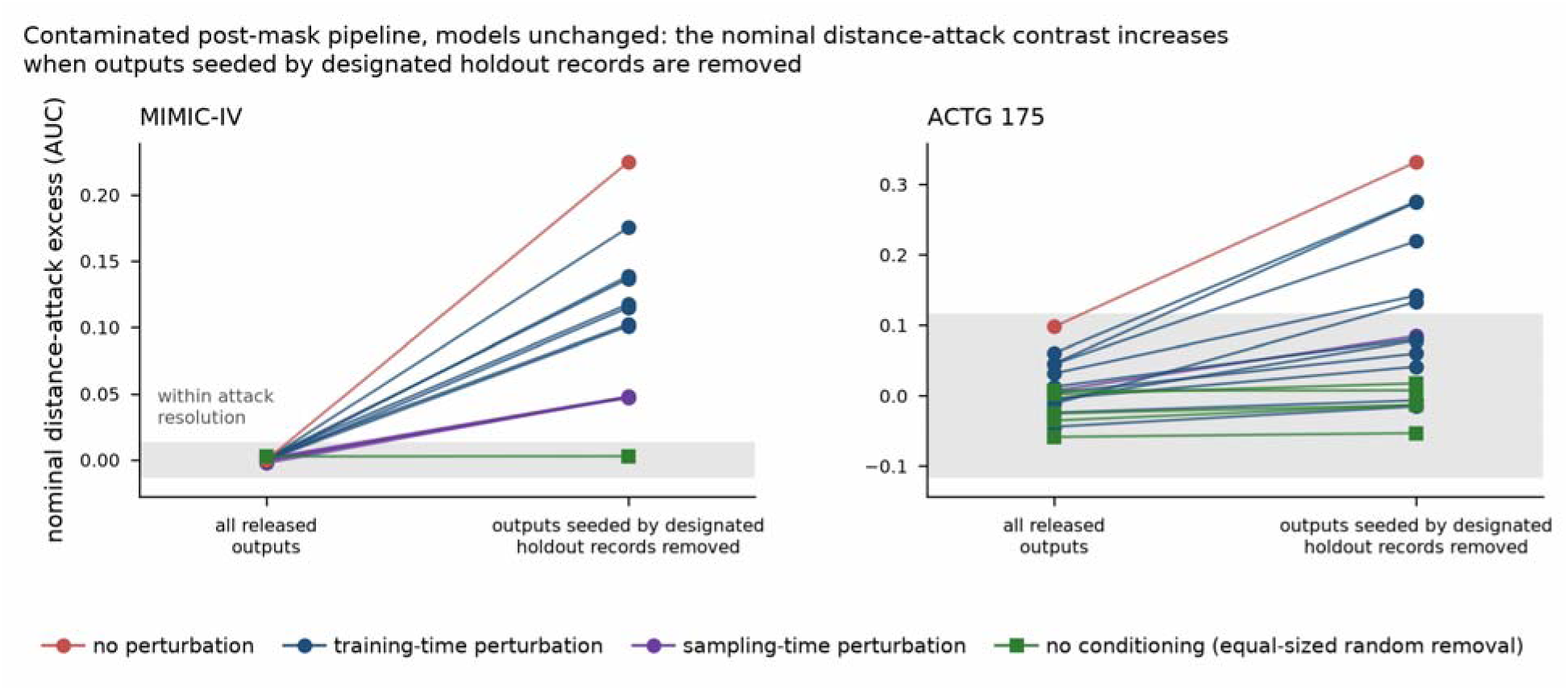
Output-removal intervention in the contaminated post-mask pipeline. Each line joins a configuration’s distance-attack excess before and after removing outputs seeded by designated holdout records. Unconditional controls use equal-sized random removal. MIMIC-IV includes 13 conditioned configurations and one unconditional output; ACTG 175 includes 13 and five. The grey band is the declared attack resolution. Models remain trained on all rows, so the post-removal contrast demonstrates sensitivity to release composition rather than the privacy of a retrained generator.

Before removal, 6 of 34 conditioned outputs exceeded the corresponding TRTR reference by more than one standard error: 4 of 14 on ACTG 175, 1 of 5 on TCGA and 1 of 15 on MIMIC-IV. None did after removal, and none of 11 unconditional outputs did before or after. TCGA lacked saved source indices, so outputs whose nearest real neighbour was a holdout row were removed instead. This proxy and the change in output composition limit a causal interpretation of the TCGA removal test. Exceeding TRTR is descriptive evidence here, not a sufficient criterion for contamination.

An earlier contaminated-pipeline comparison on TCGA gave a conditioning-associated TSTR difference of 0.032 (0.9816 versus 0.9494, nominal p < 0.001). Its five conditioned replicates varied sampling seeds from one trained model, so they do not provide five independent training replicates. The corresponding train-only comparison gave +0.0097 (p = 0.061) with additional attack signal D = 4.0. The comparisons illustrate sensitivity to membership boundaries and replication design; their difference should not be treated as a precisely isolated effect size.

### Paired retraining before and after the bundled train-only correction

With three shared seeds, the bundled train-only correction reduced TSTR and increased the relative distance-attack signal in every paired run (Figure 3, Table 6). On TCGA, TSTR changed from 0.9856 to 0.9577 (paired difference −0.0279, p = 0.008), while D changed from −0.30 to 4.00 (difference +4.30, p = 0.003). On ACTG 175, TSTR changed from 0.9666 to 0.9371 (-0.0294, p = 0.080), and D from 0.19 to 2.92 (+2.73, p = 0.001). The ACTG 175 utility reduction is directionally consistent but not significant at 0.05. These results evaluate the bundled correction in the post-mask implementation. The release size also changes from the full-table to the training-set size, so the comparison does not isolate training exposure from sample-size and other pipeline effects.

**Figure 3.**
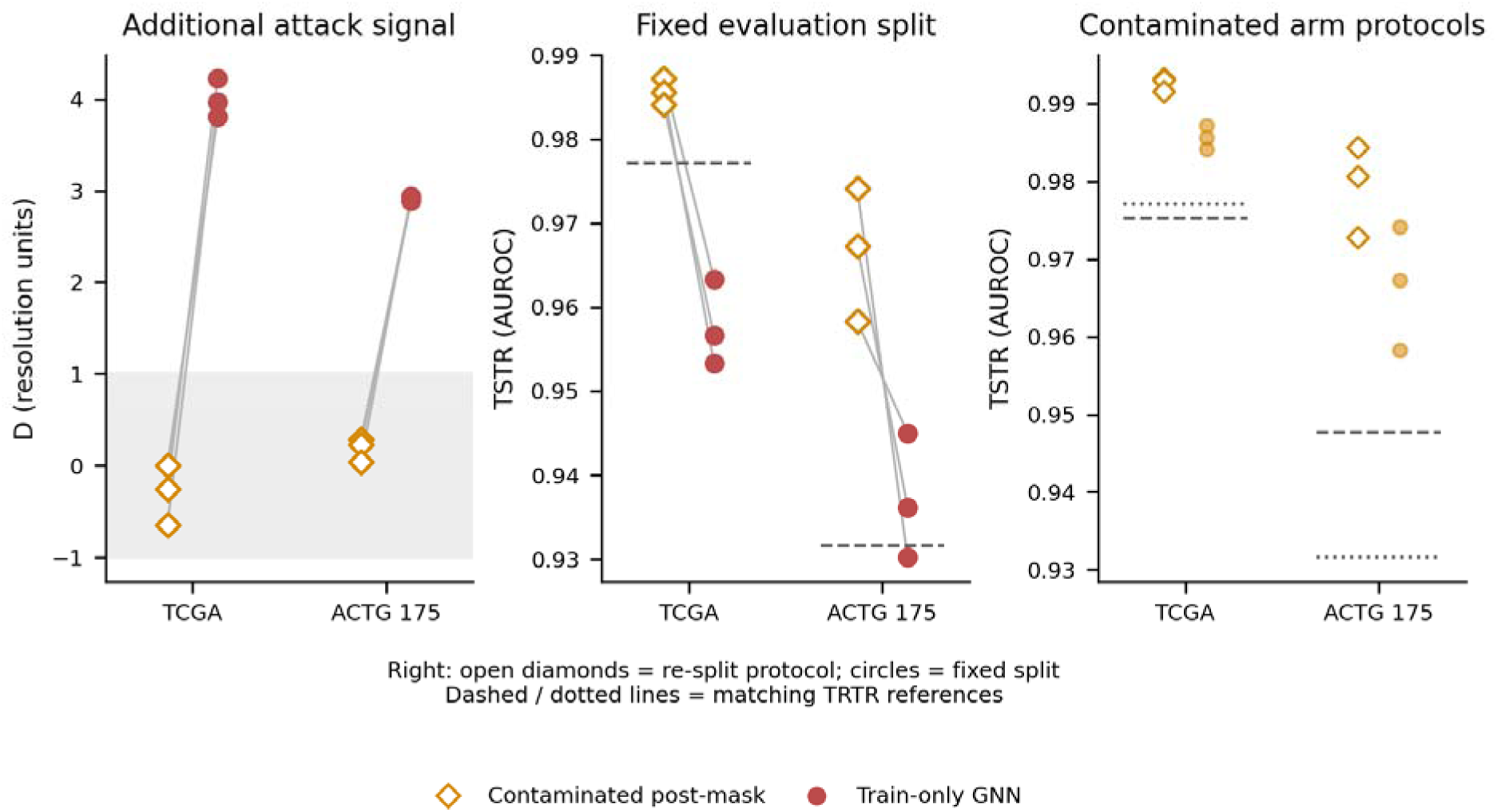
Paired contaminated post-mask and train-only GNN runs, seeds 42, 123 and 456. Left: additional distance-attack signal D relative to no conditioning. Middle: TSTR on the designated holdout, with empirical TRTR references. Right: contaminated-arm TSTR under the earlier full-table re-split protocol and fixed designated holdout, with their matching TRTR references. Lines join matching seeds in the left and middle panels. The grey band is the declared +/-1 resolutio region; it does not imply absolute privacy. No empirical reference is a theoretical ceiling.

**Table 6.**
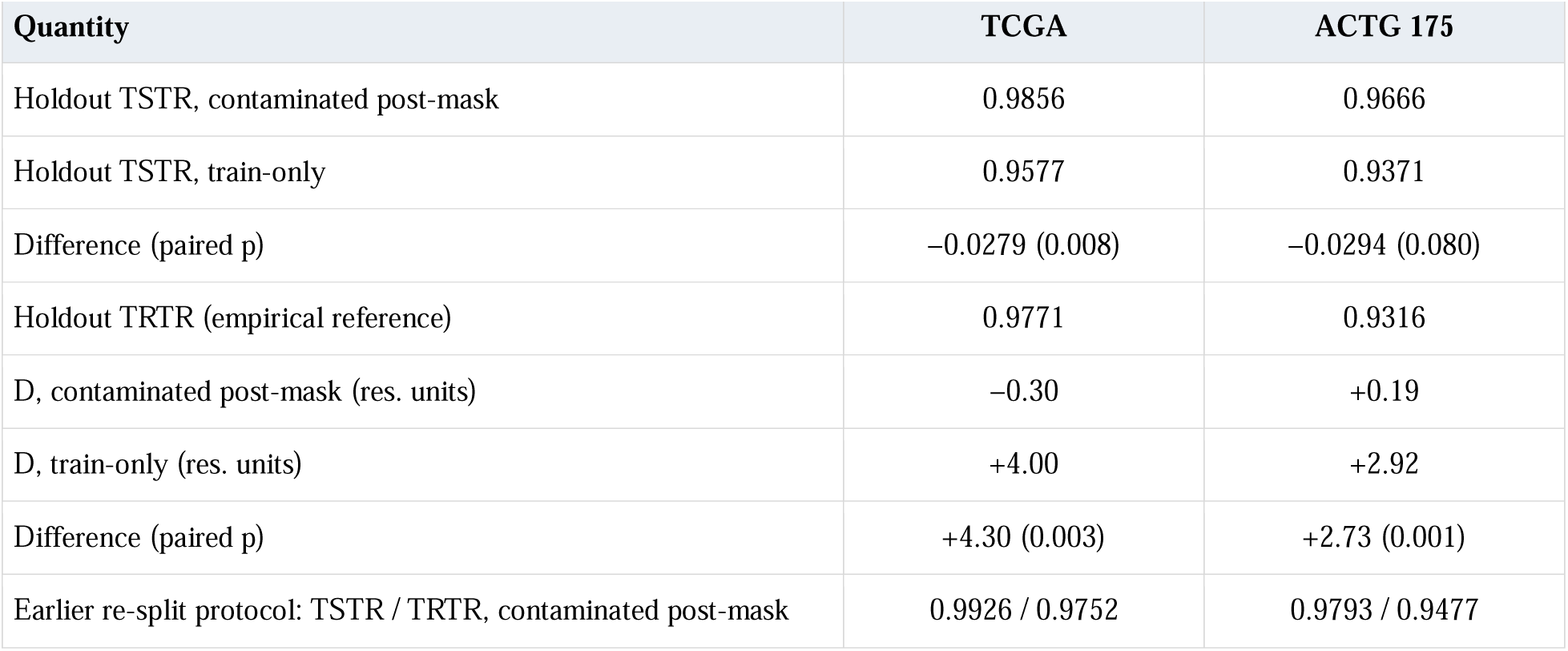
Paired contaminated post-mask and train-only GNN arms on TCGA and ACTG 175, seeds 42, 123 and 456. Means and paired t-tests are conditional on the fixed split. D is additional distance-attack signal relative to the no-conditioning backbone, in resolution units. The historical full-table re-split protocol includes generator-exposed test rows in the contaminated arm.

| **Quantity** | **TCGA** | **ACTG 175** |
|---|---|---|
| Holdout TSTR, contaminated post-mask | 0.9856 | 0.9666 |
| Holdout TSTR, train-only | 0.9577 | 0.9371 |
| Difference (paired p) | −0.0279 (0.008) | −0.0294 (0.080) |
| Holdout TRTR (empirical reference) | 0.9771 | 0.9316 |
| D, contaminated post-mask (res. units) | −0.30 | +0.19 |
| D, train-only (res. units) | +4.00 | +2.92 |
| Difference (paired p) | +4.30 (0.003) | +2.73 (0.001) |
| Earlier re-split protocol: TSTR / TRTR, contaminated post-mask | 0.9926 / 0.9752 | 0.9793 / 0.9477 |

Contaminated-arm TSTR exceeded its TRTR reference on every seed: means were 0.9856 versus 0.9771 on TCGA and 0.9666 versus 0.9316 on ACTG 175. The earlier full-table re-split protocol gave 0.9926 versus 0.9752 and 0.9793 versus 0.9477. The generator’s access to nominal test records, established independently by the loader and run evidence, explains why these are not valid estimates of out-of-training generalisation. The small nominal membership contrast is likewise uninformative because both groups were exposed.

### Testing additional MIMIC IV prediction targets

Across six non-saturated MIMIC-IV targets (TRTR 0.754-0.934), both contaminated outputs exceeded the empirical TRTR reference (Figure 4). The default output differed by +0.026 for in-hospital death, +0.038 for gender, +0.089 for insurance, +0.099 for marital status, +0.031 for admission type and +0.092 for admission location. The perturbed output differed by +0.006 to +0.035. Among 16 train-only runs, the largest positive difference on any target was +0.0011. Additional baselines ranged from 0.009 to 0.163 below TRTR. This pattern is consistent with the documented exposure of designated holdout rows; it does not make TRTR a theoretical ceiling or establish prospective clinical utility.

**Figure 4.**
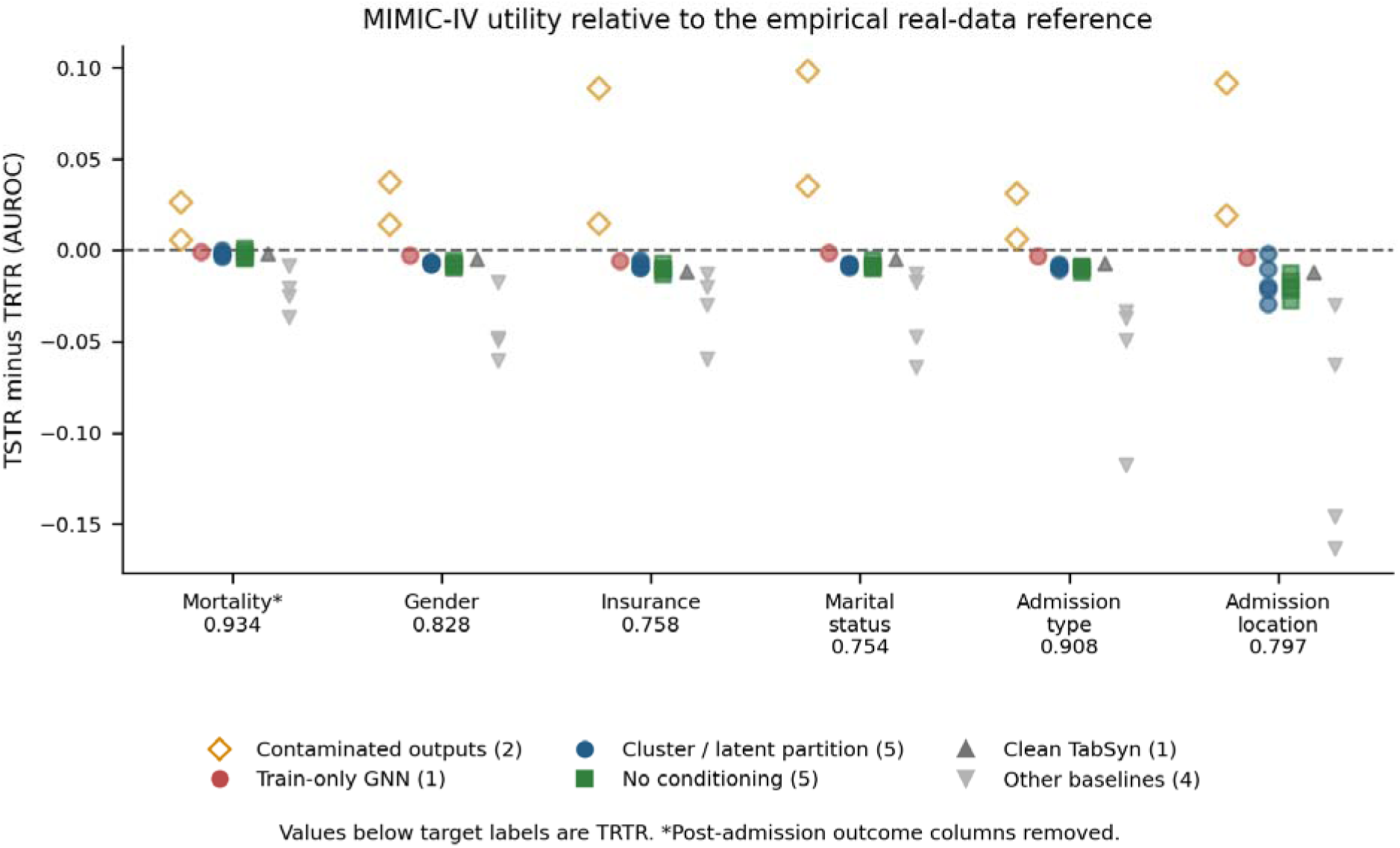
TSTR minus the empirical TRTR reference on six non-saturated MIMIC-IV tasks. Points are individual outputs or runs; contaminated outputs are shown separately from train-only methods. Both contaminated outputs exceed TRTR on all six tasks; the largest positive train-only difference is +0.0011. These are descriptive comparisons with uneve replication. For mortality, event and time_to_event_days were removed; each other target uses the remaining columns.

### Relating conditional record information to attack success

The train-only GNN-embedding arm showed additional distance-attack signal D = 4.00 on TCGA, 2.92 on ACTG 175 and 18.7 on MIMIC-IV, the cohorts with informative resolution. Across ten train-only TCGA arms including no conditioning, decodability and D were strongly associated (Spearman rho = 0.96, p < 0.001; Figure 5, Table 7). On ACTG 175 the association was weaker (rho = 0.53, p = 0.14, nine arms). GNN-condition R-squared was 0.86 on TCGA and 0.75 on ACTG 175, consistent with its training objective of predicting the record’s own latent. The association concerns the tested arms and does not by itself establish a general causal law.

**Figure 5.**
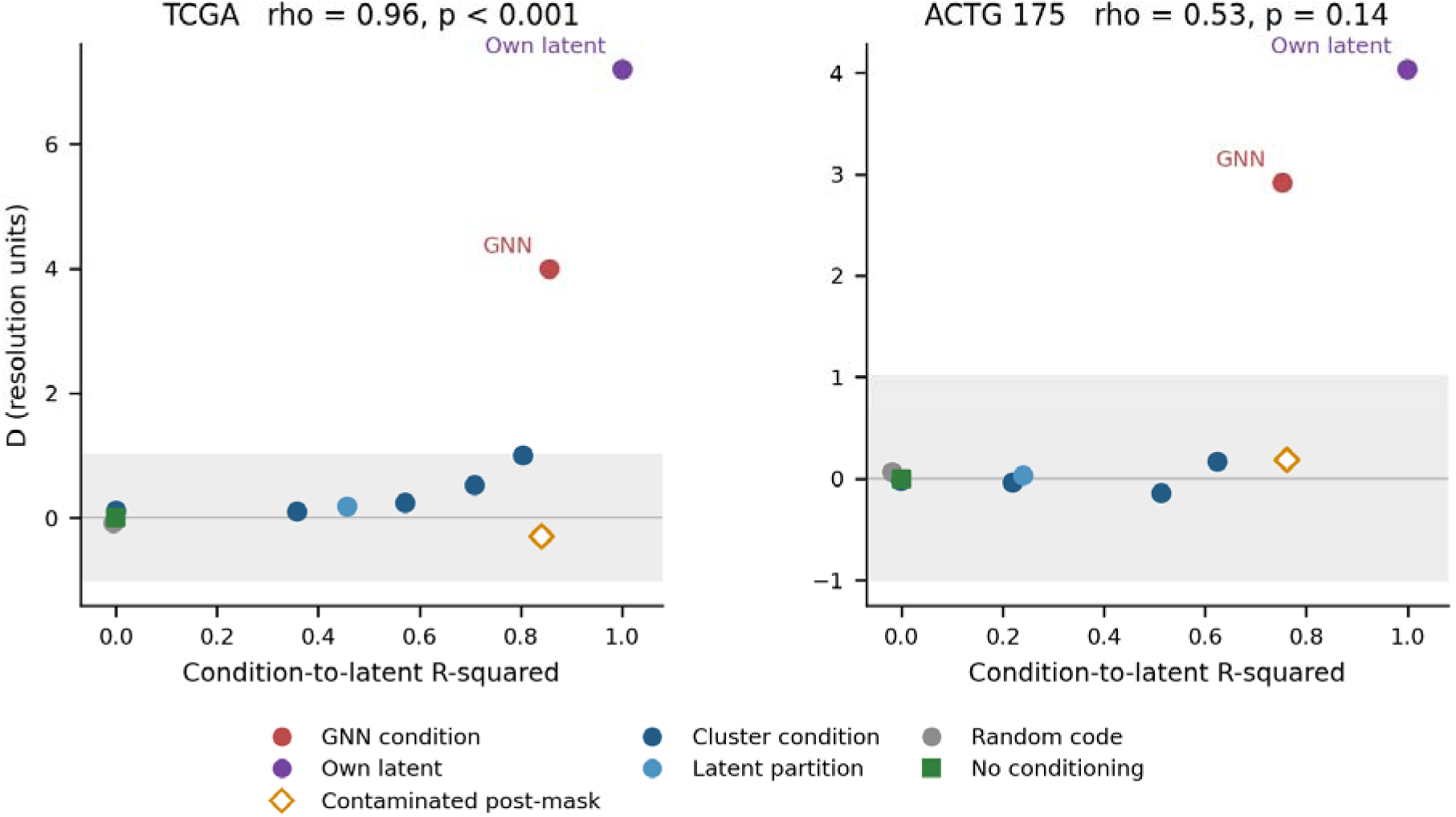
Condition-to-latent decodability and additional distance-attack signal D. R-squared is from out-of-fold ridg prediction. The association across train-only arms is strong on TCGA (rho = 0.96, p < 0.001) and weaker on ACTG 175 (rho = 0.53, p = 0.14). Random codes and record-latent conditions provide identity-content controls. Contaminated-arm points are displayed for diagnosis and excluded from the reported train-only association. The grey band marks the declare operational threshold.

**Table 7.**
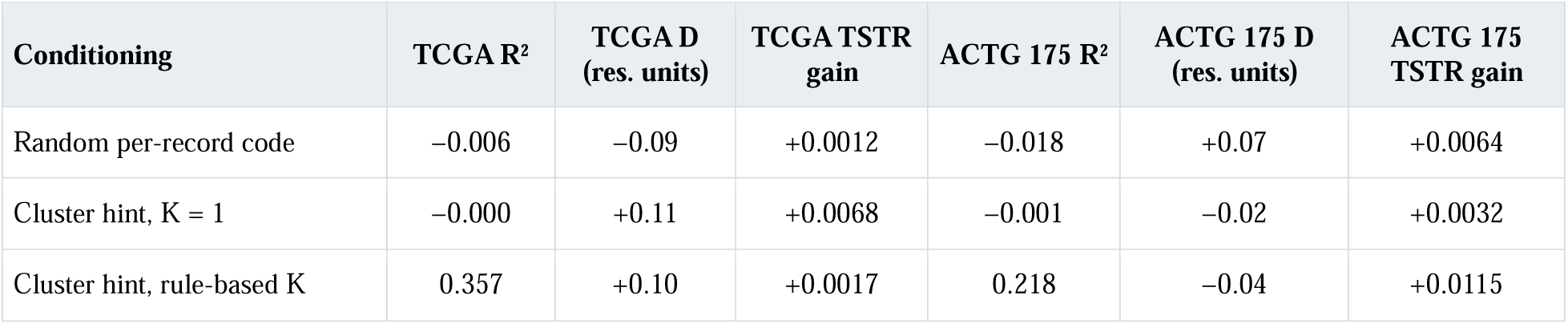
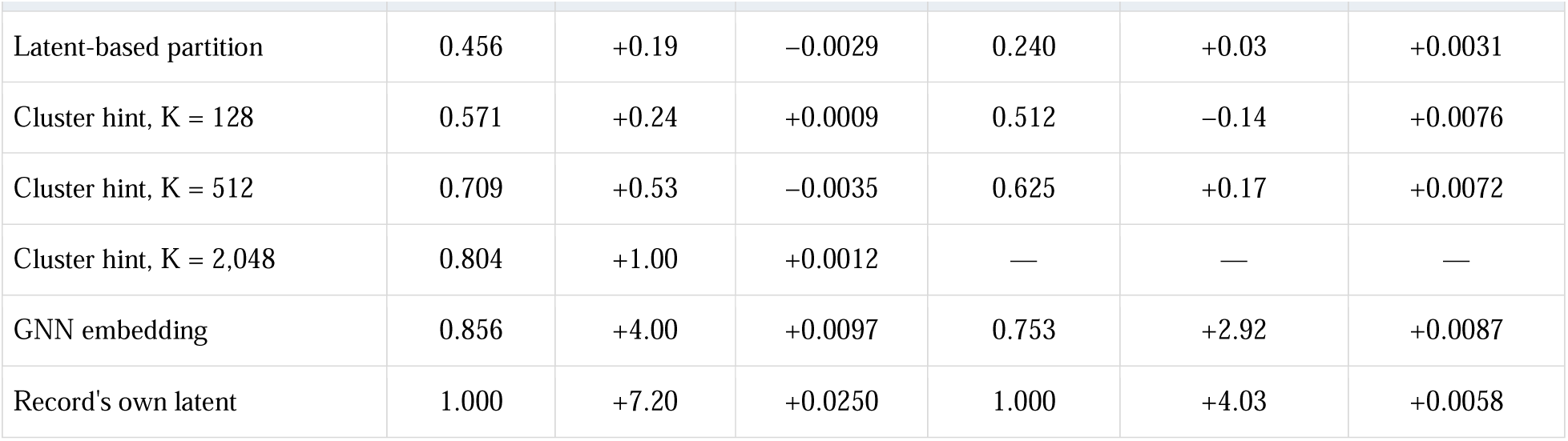
Condition decodability, additional distance-attack signal D, and utility difference from no conditioning. R-squared is out-of-fold ridge prediction of record latents. D subtracts the no-conditioning attack excess and divides by the declared resolution. Three seeds are used except K = 1, with one. Correlation including the no-conditioning reference is 0.96 on TCGA (p < 0.001, ten arms) and 0.53 on ACTG 175 (p = 0.14, nine arms). Excluding the no-conditioning reference gives 0.98 on TCGA (nine arms, p < 0.001) and 0.60 on ACTG 175 (eight arms, p = 0.12); further excluding the own-latent and random-code arms gives 0.96 (seven arms, p < 0.001) and 0.60 (six arms, p = 0.21). These are descriptive associations across experimental arms that share data and representations, not across independent patients.

Two controls distinguish a unique index from an informative condition. A random per-record code had negligible out-of-fold decodability (R-squared = −0.006 on TCGA) and no resolved additional distance-attack signal (D = −0.09). Conditioning on the record’s own latent gave the largest signal, D = 7.20 on TCGA and 4.03 on ACTG 175. Together, these controls support the role of condition content in this model family; they do not prove that arbitrary random codes can never be memorised under other training regimes.

The cluster-level experiments vary the granularity of conditioning (Figure 6). On TCGA, D increased with K (Spearman rho = 0.94, p = 0.005), reaching about 1.00 at K = 2,048 and 4.00 for one condition per record. From K = 1 to 2,048, TSTR ranged from 0.9445 to 0.9549 versus 0.9481 +/- 0.0036 without conditioning, without a monotonic trend (rho = 0.03). The K = 1 estimate uses one seed. On ACTG 175, cluster-level signals remained within the declared threshold. Its different cohort size and resolution limit cross-cohort comparisons.

On TCGA, the record-latent arm gained +0.0250 TSTR over no conditioning (two-sided p = 0.002) while also showing the largest attack signal. The GNN arm gained +0.0097 (p = 0.061); its utility difference was not significant at 0.05. The random-code arm gained +0.0012 (p = 0.62). No tested train-only TCGA arm met the prespecified combination of utility and attack-threshold criteria. Supplementary Figure S1 shows effect sizes and bootstrap intervals, which need not agree exactly with small-sample t-tests; the single-seed K = 1 estimate cannot establish a replicated advantage.

### Checking a second attack and its sampling sensitivity

MIA-EPT detected the train-only GNN arm on TCGA (excess 0.0759; resolution 0.0469) and ACTG 175 (0.2086; resolution 0.1097); no-conditioning and rule-based cluster arms remained within resolution (Table 8). On MIMIC-IV, the initial 20,000-row predictor cap yielded excess −0.0121. The member selection and subsampling procedures shared a seeded random sequence: the copy control retained all selected members, whereas only 14.1% of source-linked synthetic counterparts were retained in the GNN arm, close to the 14.2% fair-sampling share. The copy control therefore did not establish comparable sensitivity for that arm. A declared full-table rerun yielded excess 0.0664, or 5.18 resolution units, while no conditioning remained at −0.17 units and the copy control at 38.4. This supports a sensitivity limitation of the capped evaluation and corroborates disclosure in the tested GNN run. The full-table comparison uses one generator seed per arm.

**Table 8.** MIA-EPT results for train-only arms. Generator seed is 42 except MIMIC-IV no conditioning, which uses 123. Excess is attack AUC minus negative-control AUC; resolutions are 0.0469, 0.1097 and 0.0128. Capped and full-table MIMIC-IV evaluations are distinguished because their effective access to source-linked records differs. These are attack-specific results, not privacy guarantees.

| Cohort | Arm | Attack AUC | Control AUC | Excess | × resolution |
|---|---|---|---|---|---|
| TCGA | GNN embedding | 0.5623 | 0.4864 | +0.0759 | +1.62 |
| TCGA | No conditioning | 0.5218 | 0.4916 | +0.0302 | +0.64 |
| TCGA | Cluster hint, rule-based K | 0.4755 | 0.4640 | +0.0115 | +0.25 |
| TCGA | Verbatim copy | 0.8927 | 0.5027 | +0.3900 | +8.32 |
| ACTG 175 | GNN embedding | 0.7036 | 0.4950 | +0.2086 | +1.90 |
| ACTG 175 | No conditioning | 0.6415 | 0.5528 | +0.0887 | +0.81 |
| ACTG 175 | Cluster hint, rule-based K | 0.5605 | 0.5718 | −0.0113 | −0.10 |
| ACTG 175 | Verbatim copy | 0.9896 | 0.4369 | +0.5527 | +5.04 |
| MIMIC-IV, subsampled predictors | GNN embedding | 0.4965 | 0.5086 | −0.0121 | −0.94 |
| MIMIC-IV, subsampled predictors | No conditioning | 0.4980 | 0.5076 | −0.0096 | −0.75 |
| MIMIC-IV, subsampled predictors | Verbatim copy | 0.9998 | 0.5019 | +0.4978 | +38.89 |
| MIMIC-IV, full synthetic table | GNN embedding | 0.5694 | 0.5031 | +0.0664 | +5.18 |
| MIMIC-IV, full synthetic table | No conditioning | 0.4969 | 0.4992 | −0.0022 | −0.17 |
| MIMIC-IV, full synthetic table | Verbatim copy | 0.9997 | 0.5084 | +0.4913 | +38.38 |

### Evaluating utility after restricting the conditioning channel

Among comparisons with adequate replication and informative attack calibration, no tested train-only variant with attack signals within the declared thresholds showed a significant TSTR gain over clean TabSyn (Figure 7, Table 9). The ACTG 175 rule-based cluster arm differed by +0.0058 (p = 0.45). WBCD’s largest difference was +0.0080 (p = 0.11), but privacy could not be assessed there. On TCGA, the GNN arm differed by +0.0077 (p = 0.12; D = 4.0), and the record-latent arm by +0.0230 (p = 0.006; D = 7.2). Contaminated-arm differences were +0.0356 on TCGA and +0.0323 on ACTG 175 and are not valid train-only improvements. None of 18 fidelity comparisons reached p < 0.05; the smallest was 0.056 for the ACTG 175 GNN arm. These null comparisons leave smaller effects unresolved.

**Figure 6.**
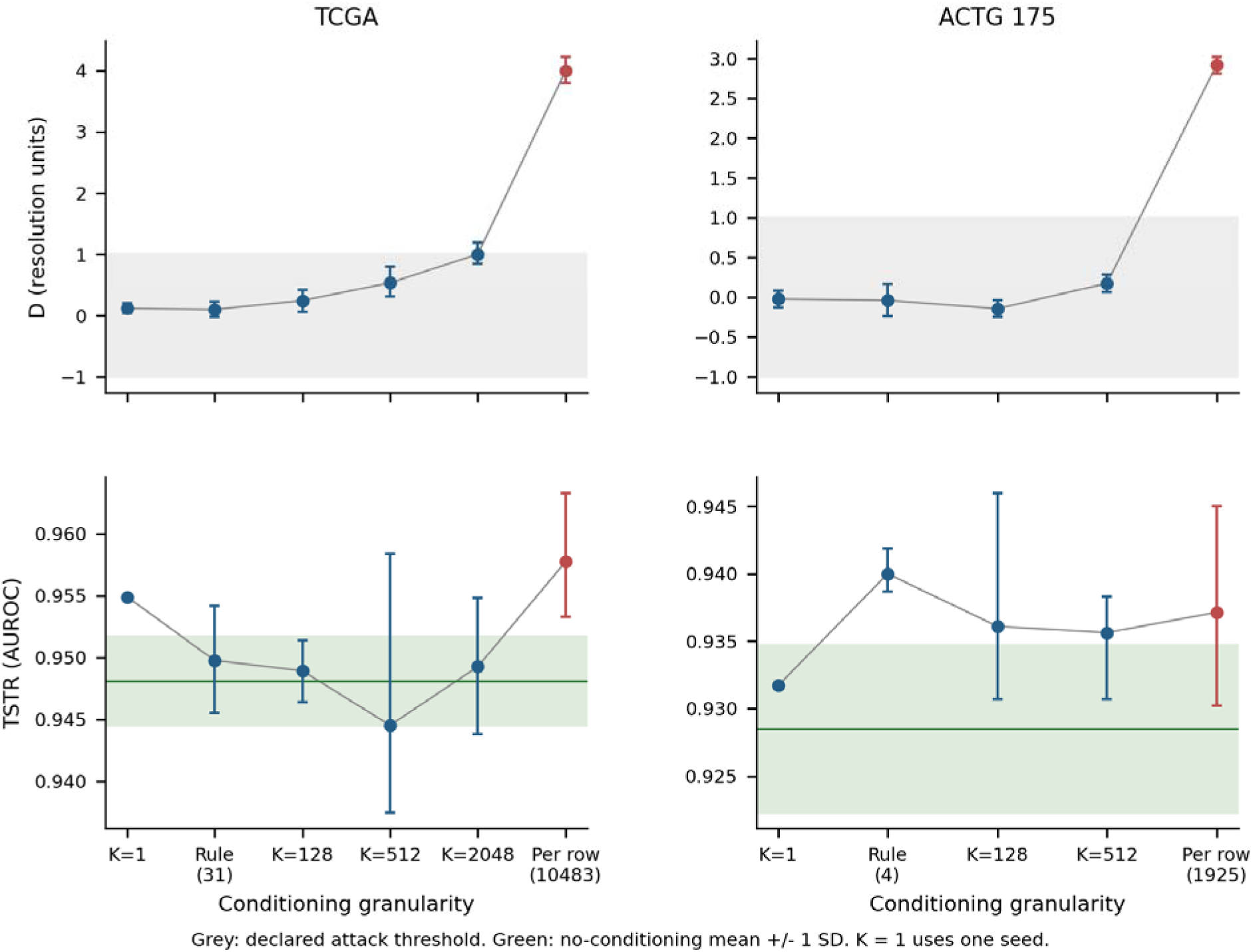
Conditioning granularity in train-only arms. Points show arm means and bars show the reported seed-bootstrap 95% intervals. The upper panels display D; lower panels display TSTR with the no-conditioning mean +/-1 SD. TCGA shows increasing attack signal with granularity without a monotonic utility increase. ACTG 175 cluster-level signals remain within threshold. K = 1 has one seed and no between-seed uncertainty; an attack-difference interval can still reflect the reference arm. Connecting lines guide the eye, not a fitted continuous dose-response model.

**Figure 7.**
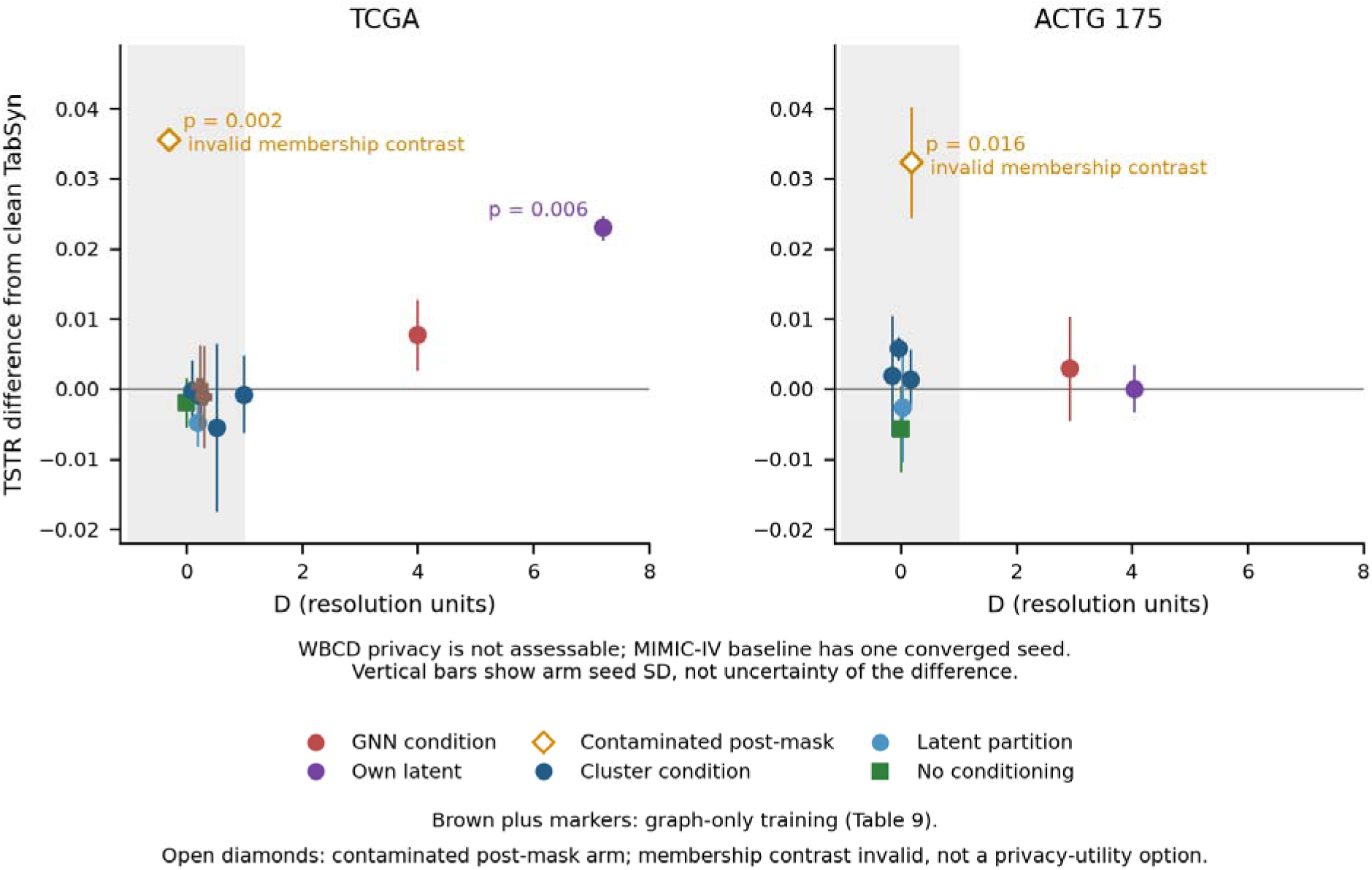
Utility difference from clean TabSyn and additional distance-attack signal D on TCGA and ACTG 175. Points are arm means and vertical bars are generator-seed SD, not confidence intervals for the difference between arms. Annotated p-values are the nominal Welch tests reported in Table 9. The threshold band describes additional signal relative to no conditioning, not absence of absolute leakage. WBCD is excluded from the joint plot because its privacy coordinate is not assessable; its utility results remain in Table 9. MIMIC-IV lacks a replicated clean TabSyn comparator and is reported descriptively in Figure 4 and Table 9. Open diamonds mark the contaminated post-mask arm; its nominal membership contrast is invalid, so it is shown for reference and is not a privacy–utility option.

**Table 9.** Utility comparisons with the corrected local TabSyn comparator. Welch tests use generator seeds. MIMIC-IV has one converged clean TabSyn seed and therefore no inferential comparison; its main mortality task is saturated (Figure 4 reports additional tasks). D is relative to the no-conditioning backbone. WBCD attack interpretation is not assessable. Nominal p-values and non-significant differences do not establish equivalence.

| Cohort | Arm | n | Holdout TSTR | Difference vs clean TabSyn | Welch p | D<br>(res. units) |
|---|---|---|---|---|---|---|
| TCGA | Clean TabSyn (reference) | 3 | 0.9500 | 0 (reference) | — | +0.26 |
| TCGA | No conditioning | 3 | 0.9481 | −0.0020 | 0.586 | +0.00 |
| TCGA | GNN embedding | 3 | 0.9577 | +0.0077 | 0.122 | +4.00 |
| TCGA | Cluster hint, rule-based K | 3 | 0.9498 | −0.0003 | 0.945 | +0.10 |
| TCGA | Latent-based partition | 3 | 0.9452 | −0.0048 | 0.217 | +0.19 |
| TCGA | Cluster hint, K = 128 | 3 | 0.9489 | −0.0011 | 0.735 | +0.24 |
| TCGA | Cluster hint, K = 512 | 3 | 0.9445 | −0.0055 | 0.520 | +0.53 |
| TCGA | Cluster hint, K = 2,048 | 3 | 0.9493 | −0.0008 | 0.861 | +1.00 |
| TCGA | Record's own latent | 3 | 0.9730 | +0.0230 | 0.006 | +7.20 |
| TCGA | Contaminated post-mask | 3 | 0.9856 | +0.0356 | 0.002 | −0.30 |
| TCGA | Graph only during training, p = 0.5 | 3 | 0.9503 | +0.0003 | 0.948 | +0.24 |
| TCGA | Graph only during training, p = 1.0 | 3 | 0.9488 | −0.0012 | 0.818 | +0.30 |
| ACTG 175 | Clean TabSyn (reference) | 3 | 0.9342 | 0 (reference) | — | −0.05 |
| ACTG 175 | No conditioning | 3 | 0.9285 | −0.0058 | 0.471 | +0.00 |
| ACTG 175 | GNN embedding | 3 | 0.9371 | +0.0029 | 0.720 | +2.92 |
| ACTG 175 | Cluster hint, rule-based K | 3 | 0.9400 | +0.0058 | 0.446 | −0.04 |
| ACTG 175 | Latent-based partition | 3 | 0.9316 | −0.0026 | 0.747 | +0.03 |
| ACTG 175 | Cluster hint, K = 128 | 3 | 0.9361 | +0.0019 | 0.825 | −0.14 |
| ACTG 175 | Cluster hint, K = 512 | 3 | 0.9356 | +0.0014 | 0.847 | +0.17 |
| ACTG 175 | Record's own latent | 3 | 0.9342 | 0.0000 | 1.000 | +4.03 |
| ACTG 175 | Contaminated post-mask | 3 | 0.9666 | +0.0323 | 0.016 | +0.19 |
| WBCD | Clean TabSyn (reference) | 3 | 0.9830 | 0 (reference) | — | not assessable |
| WBCD | No conditioning | 3 | 0.9885 | +0.0055 | 0.217 | not assessable |
| WBCD | GNN embedding | 3 | 0.9903 | +0.0073 | 0.133 | not assessable |
| WBCD | Cluster hint, rule-based K | 3 | 0.9910 | +0.0080 | 0.108 | not assessable |
| WBCD | Latent-based partition | 3 | 0.9874 | +0.0044 | 0.341 | not assessable |
| MIMIC-IV | Clean TabSyn (reference) | 1 | 0.9979 | 0 (reference) | — | +0.07 |
| MIMIC-IV | No conditioning | 2 | 0.9978 | −0.0001 | — | +0.00 |
| MIMIC-IV | GNN embedding | 1 | 0.9977 | −0.0002 | — | +18.65 |
| MIMIC-IV | Latent-based partition | 1 | 0.9978 | −0.0001 | — | −0.09 |
| MIMIC-IV | Cluster hint, rule-based K (clipped) | 3 | 0.9978 | −0.0001 | — | +0.07 |
| MIMIC-IV | No conditioning (clipped) | 3 | 0.9978 | −0.0001 | — | +0.04 |

Neighbour mixup during training also failed to meet its declared utility criterion on TCGA. TSTR differences from clean TabSyn were +0.0003 at probability 0.5 (p = 0.95) and −0.0012 at probability 1.0 (p = 0.82). At probability 1.0, fidelity worsened (KS +0.0126, p = 0.001; correlation error +0.0109, p = 0.005). No additional attack-signal difference was resolved. A zero-mixup control matched clean TabSyn metric for metric. The prespecified gain of at least 0.010 at p < 0.025 was not achieved; this tests two settings in one cohort, not every use of graph information during training.

Clean TabSyn’s mean TSTR was already within 0.004 of TRTR on ACTG 175, WBCD and the saturated MIMIC-IV target, while the TCGA gap was 0.027. Its holdout KS was 0.004-0.015 above that of a verbatim-copy reference. These comparisons describe the benchmark’s dynamic range for the chosen metrics. Neither TRTR nor a copy-control fidelity score defines a universal optimum.

ACTG 175 provides a useful comparator-dependent exception. The rule-based cluster arm and K = 512 gained +0.0115 and +0.0072 over the no-conditioning backbone, with seed-bootstrap intervals excluding zero and additional attack signals within threshold. Against clean TabSyn, the differences were +0.0058 (p = 0.45) and +0.0014 (p = 0.85). With 214 holdout rows and three seeds, these observations motivate cautious reporting rather than a claim that conditioning never helps.

## 5 Discussion

### Principal findings

This case study identifies two interacting evaluation mechanisms and one design-specific disclosure channel. First, nominal holdout exposure compromises both generalisation testing and the definition of non-members. Second, source-linked outputs for both nominal groups can suppress the distance contrast even when outputs remain close to their source records. Third, after the training boundary is enforced, informative per-record conditions are associated with measurable membership-attack signals. Output removal, paired retraining, identity-content controls and a second attack family provide complementary evidence. The contribution is a diagnostic sequence for separating these mechanisms within the studied implementation.

### Interpreting the diagnostic interventions

The interventions answer different questions. Output removal changes which synthetic counterparts are released while holding the trained model fixed; the resulting attack change supports an output-composition mechanism. Paired train-only retraining changes the training boundary and associated run controls; it shows that the contaminated evaluation overstated utility in these runs while obscuring the residual attack signal. The random-code and record-latent controls then test whether a unique per-record condition or its recoverable content better explains that signal. Treating these comparisons separately avoids attributing every historical change to a single repair.

The released feature mask is a related implementation finding. It prevented the inspected code from using patient-specific graph inputs, so conclusions about similarity conditioning require an executable state in which that information is present. The historical mask repair coincided with other training changes and was not re-run here. We therefore distinguish the documented defect from any claim about its isolated quantitative effect or the exact origin of each published result.

### Why contamination affects utility and membership contrasts

A privacy audit needs a valid distinction between exposed and unexposed records. When both nominal groups enter training and supply conditions, comparable proximity to synthetic outputs can make the contrast small. A near-null AUC in that setting is not evidence of privacy. At the same time, training exposure of evaluation records can improve TSTR without demonstrating out-of-training generalisation. The observed effects need not occur with equal strength in every generator; here, provenance checks and controlled interventions show that they matter in the post-mask pipeline.

TSTR above TRTR is a reason to inspect an evaluation, not proof of contamination. TRTR measures a particular classifier trained on a finite sample. A synthesis procedure can alter smoothing, weighting or regularisation, so a TSTR gain does not necessarily require access to test records. In this study, the contamination diagnosis instead rests on actual loader behaviour, saved-run evidence and the response to interventions. The MIMIC-IV pattern across six tasks provides additional consistency evidence. Small-holdout seed variation further limits the heuristic: clean TabSyn on ACTG 175 ranged from −0.008 to +0.013 around TRTR.

### Record content as a conditioning channel

The GNN condition carries information about a real record beyond the randomly sampled diffusion noise. Its training target is the record’s latent, and its inputs include that record’s features. High out-of-fold decodability and the record-latent control support a content-mediated disclosure mechanism. The random-code control shows that uniqueness alone did not produce the same signal under the tested training regime. Cluster-level sampling reduced the additional signal below the operational threshold in several arms, but the present comparisons did not establish a reliable utility advantage over clean TabSyn. This supports a bounded trade-off result rather than the identity of all utility gains with leakage.

The findings apply to the tested single-table design, conditioning strategies and attacks. They do not show that graph structure is generally unhelpful, that all record-specific conditions are unsafe, or that cluster-level conditions are private against every adversary. Relational generation with genuine links between tables and a separately sampled relational structure is a different setting [18].

### Controls matched to the failure mechanism

Different checks are needed at different boundaries. Input-variation and conditioning-ablation checks establish whether the intended information path is active. Saved row indices and latent provenance establish which records each stage used. Copy and non-member controls test the behaviour of the attack, while sampling-overlap checks test whether those controls represent the evaluated release. The MIMIC-IV MIA-EPT result illustrates the last distinction: a positive control can be easy to detect because its selected members are overrepresented in the predictor-training sample. A successful control therefore needs to be interpreted alongside the sampling procedure and the threat model.

### Implications for evaluation practice

The safeguards below turn these observations into reproducible checks. Their purpose is to validate the information boundary and the sensitivity of the measurements before interpreting a utility-privacy trade-off. Detection scores remain useful for distinguishability, and record-matching measures can reveal near-copying, but neither alone establishes membership privacy [11,12]. When the original privacy evidence does not support its interpretation, that limitation should be stated explicitly and the affected claim reassessed.

### Evaluation safeguards

1. Verify which rows every stage reads, including the encoder, graph, condition pool, model selection and generator. Preserve indices or provenance checks with each run.
2. Keep an independent evaluation set outside training and model selection. Do not infer independence merely from a stored split name.
3. Report TSTR beside a matched TRTR reference and its uncertainty. Investigate unexpected gains using provenance and controls rather than treating TRTR as an upper bound.
4. Calibrate each attack on a copy control and an appropriate non-member control. Report raw AUC, control AUC, excess, and any baseline-relative transformation separately.
5. Check whether control construction, subsampling and source-linked overlap give the control a different exposure pattern from the evaluated arm.
6. Measure what record information a condition encodes, using identity-content controls and reconstruction or decodability probes appropriate to the threat model.
7. Verify adapted baselines against their intended training protocol. Match splits and replication where feasible and report divergence rather than silently substituting runs.
8. Check that conditions vary across records and that ablation changes the intended information path. A code repair and its isolated empirical effect are separate questions.
9. Pair fidelity improvements with record-matching and attack checks. Neither high fidelity nor novelty alone establishes privacy.
10. Record decision rules and deviations with their timing. Keep negative results bounded by the tested methods, attacks, datasets and statistical power.

### Limitations

Most arms have only three seeds on one fixed split and share fitted representations; seed-bootstrap intervals are coarse, non-significant tests do not establish equivalence, and excluded divergence limits comparison to converged runs. MIMIC-IV clean TabSyn has one converged seed, as do its GNN arm and full-data MIA-EPT comparison. WBCD does not provide adequate attack resolution. The MIMIC-IV cohort keeps one admission per patient and requires complete laboratory panels, so it does not represent all hospital admissions. Several utility targets are descriptive rather than prospective clinical prediction tasks. The distance-attack resolution is an approximate null calibration and baseline subtraction is not a confidence bound for absence of leakage. Attack adaptations, ordered holdout subsets and subsampling choices also limit generalisation. The latent-partition control retains GNN-derived conditions and does not isolate all effects of graph encoding. We studied one implementation family and two attack families, without a formal guarantee such as differential privacy. The paired pipeline correction bundles changes to training membership, run controls and release size; the mask repair was not isolated experimentally. Historical working-record values and the initial preprint’s exact numbers were not regenerated. Output-file size alone does not determine training exposure, and the association between released code and every original published result remains incomplete. Output removal tests a contaminated trained model and cannot bound the privacy of a separately retrained one.

## 6 Conclusion

Within the tested TabGraphSyn pipeline, designated holdout exposure inflated utility estimates and weakened nominal membership contrasts. After restricting training to training rows, informative per-record conditions retained measurable disclosure signals, supported by content controls and a second attack family. Several cluster-level alternatives reduced the additional signal within the declared thresholds, but a reliable utility advantage over clean TabSyn was not established. The practical contribution is a mechanism-based evaluation sequence: verify training membership, validate attack sensitivity under the actual sampling procedure, and test what record information the condition carries. These checks support narrower and more defensible claims about synthetic clinical data.

## Data availability

TCGA clinical data are available through the UCSC Xena platform [21]. ACTG 175 [22] and the Wisconsin Breast Cancer (Original) dataset [23] are publicly available. MIMIC-IV is available to credentialed users through PhysioNet under its data-use agreement [24]; we release no MIMIC-IV-derived records. Dated experiment declarations, aggregate result tables, figure source tables and SHA-256 manifests are provided as Supplementary Data S1, whose README identifies the included supporting files.

## Code availability

The full source code is maintained in a private laboratory repository and is not publicly available. Aggregate results and supporting documentation are provided in Supplementary Data S1.

## Ethics statement

This study included a secondary analysis of de-identified MIMIC-IV data accessed through PhysioNet under its credentialing requirements and data use agreement. Ethical oversight of the original MIMIC-IV research resource was provided by the Institutional Review Board at Beth Israel Deaconess Medical Center, which authorized data sharing and waived individual informed consent [24]. The present MIMIC-IV analyses involved no new participant recruitment or direct contact with patients.

## Use of generative AI

Generative AI tools from Anthropic (Claude, including Claude Science) and OpenAI (ChatGPT and Codex) were used to assist with code development and debugging, execution of computational experiments, and manuscript revision. Before submission of this preprint, the authors manually reviewed all content, including AI-assisted code, analyses, results, and manuscript text. The authors take full responsibility for the accuracy, integrity, and originality of the work.

## Data Availability

TCGA clinical data are available through the UCSC Xena platform. ACTG 175 and the Wisconsin Breast Cancer (Original) dataset are publicly available. MIMIC-IV is available to credentialed users through PhysioNet under its data-use agreement; we release no MIMIC-IV-derived records. Dated experiment declarations, aggregate result tables, figure source tables and SHA-256 manifests are provided as Supplementary Data S1, whose README identifies the included supporting files. The full source code is maintained in a private laboratory repository and is not publicly available.

https://archive.ics.uci.edu/dataset/890/aids+clinical+trials+group+study+175

https://archive.ics.uci.edu/dataset/15/breast+cancer+wisconsin+original

https://xenabrowser.net/datapages/?dataset=Survival_SupplementalTable_S1_20171025_xena_sp&host=https%3A%2F%2Fpancanatlas.xenahubs.net&removeHub=https%3A%2F%2Fxena.treehouse.gi.ucsc.edu%3A443

https://physionet.org/content/mimiciv/

## Figures

Figures 3-7 and Supplementary Figure S1 use the same aggregate source tables as the reported analyses. D denotes additional distance-attack signal relative to no conditioning, in the declared resolution units. Shaded threshold regions are operational decision regions, not privacy guarantees.

## Appendix A Relationship to the initial preprint and reporting corrections

**Table A1.** Scope of the current evidence for claims in the initial preprint [1]. Implementation findings, results from later variants, and unreproduced historical values are distinguished. A claim requiring re-evaluation is not necessarily numerically disproved.

| **Claim in version 1** | **Status** |
|---|---|
| The diffusion model is guided by GNN embeddings of a patient-similarity graph (title, abstract and Methods) | The inspected released code masks the single table and removes patient-specific graph information. Later post-mask variants do use informative conditions. Mapping each original published result to its executable state remains incomplete. |
| Classifiers trained on synthetic data matched real-data performance (WBCD AUC 0.9938 vs 0.9933 for real data) | Requires re-evaluation with verified training-only inputs and an independent test. The specific WBCD number was not regenerated. Paired later-pipeline experiments quantify contamination effects on TCGA and ACTG 175, not that historical WBCD estimate. |
| Graph embeddings yielded up to 11% improvement in ten-fold augmentation; removing the GNN degrades fidelity | The inspected masked implementation cannot establish a benefit from patient-specific similarity information. The historical augmentation experiment was not rerun. Tested train-only variants did not establish a reliable gain over clean TabSyn; this does not disprove every graph-based augmentation approach. |
| Up to 3.5% lower marginal and 2.61% lower pairwise-correlation error than the best baseline | Not independently reproduced. The current 18 fidelity comparisons with clean TabSyn found no significant differences, which does not establish equivalence or reconstruct the historical rankings against every baseline. |
| Privacy: Detection Score and DCR audits indicate robust de-identification | The original evidence does not establish robust de-identification: detection measures distinguishability and DCR alone does not establish membership privacy. Later train-only per-record variants show attack-detectable disclosure. A privacy claim must be tied to a verified implementation, threat model and evaluation. |
| Survival analysis: synthetic data recover significant Cox covariates (F1 0.857) | Not re-examined. No conclusion about validity of the F1 = 0.857 result follows from the present classification analyses. The reporting discrepancies below require correction, and survival-specific validation remains separate. |
| 100% data validity | The audit does not reassess this reported value. Type or schema validity is distinct from clinical plausibility, utility and privacy. |
| "The models were only trained on training set so that holdout set is hidden from model" (Privacy Preservation section) | The inspected loader and available later-run evidence contradict this statement for those implementations. Provenance of individual original outputs requires separate confirmation; a nominal holdout declaration alone is insufficient. |

**Table A2.** Reporting corrections and interpretation checks in the initial preprint. Arithmetic and text-table discrepancies can be checked independently of the mechanism experiments. Where an explanation for a discrepancy is unknown, no attribution of intent or copying is made.

| Location in version 1 | As published | Correct or consistent value | Effect |
|---|---|---|---|
| Privacy section, Table 6 (Detection Score), Avg. column | TabGraphSyn 91.95%; all six averages | Means of the three cohort values: TabGraphSyn 84.30%, TabSyn 84.81%, TabDDPM 76.11%, TabDiff 66.33%, CTGAN 37.76%, CTAB-GAN+ 8.54% | All six published averages differ from the means of the displayed cohort values. The published averages already ranked TabSyn (92.45%) above TabGraphSyn (91.95%); recomputation does not change that order. |
| WBCD classification paragraph | TabGraphSyn F1 98.83%, AUC 99.96% | Table 5: F1 0.9768, AUC 0.9938. F1 0.9883 is TabDDPM's; 0.9996 appears nowhere | Text contradicts its own table; TabDDPM has the best F1 and AUC. |
| Table 5 (TSTR, WBCD) | Real data F1 0.9588 | Five of six generators have F1 above real data | TSTR above TRTR is a descriptive observation and a reason to verify the evaluation split; it does not by itself establish contamination. |
| Table B1, TabDiff column | TP 3, FP 1, FN 5, TN 4; F1 0.50 (identical to CTGAN) | From the check marks: TP 3, FP 0, FN 5, TN 5; precision 1.00, recall 0.375, F1 0.545 | The published counts duplicate the CTGAN counts despite different marks. The cause is unverified. Corrected F1 ranks TabDiff above CTGAN. Recomputed from the table's own marks; the Cox models were not rerun. |
| Table B1, TabDDPM column | TN 0 | TN 2 (13 variables; TP 7, FP 3, FN 1) | Counts did not sum to 13; F1 unchanged. Recomputed from the table's own marks; the Cox models were not rerun. |
| Ablation paragraph | "Removal of graph embeddings reduced marginal distribution errors by ~11.21% and pairwise errors by ~10.54%" | Adding graph embeddings reduced errors by those amounts, at r = 10 only | Direction inverted. At r = 0.1 the graph increased marginal error on TCGA and pairwise error on AIDS (also at r = 0.5). |
| Tables 8–9, Improv. rows | Relative reduction of the summed error across cohorts | Alternative aggregation, mean of per-cohort reductions: marginal 0.69–9.72%, pairwise −0.01–8.91% | Neither aggregation is incorrect; they weight cohorts differently. The aggregation used should be stated. |
| Dataset section | "WBCD (Wisconsin Breast Cancer Diagnostic) dataset" | Wisconsin Breast Cancer (Original): 683 complete records, 9 integer features (Table 1 of version 1 and reference 15) | Dataset misnamed; the Diagnostic dataset has 569 records and 30 real-valued features. |
| Table 2, TabGraphSyn Avg. | 2.57 | 2.58 when computed from the displayed rounded values 1.42, 3.94 and 2.37 | Possible rounding discrepancy; without the unrounded values an error in the published mean is not established. |
| Conclusion, Table 10 description | "The Avg Rank column reports the mean rank" | The table reports the sum of ranks | Description does not match the table. |
| Privacy section, Equation 6 and text | A Detection Score near 1 "suggests higher privacy" | A verbatim copy of the training data scores 1 by construction | The metric measures distinguishability, not privacy. |
| DCR paragraph | Ideal value 50% | A 50% reference depends on the distance definition, reference-set sizes and sampling; it is not a universal privacy optimum. | The appropriate reference depends on train/holdout sizes and sampling. The reported DCR values alone do not establish privacy. |
| Implementation Details | "we set the K-NN graph parameter k = 3" | The released code fixes k at 5 (k = min(5, n − 1)) and builds the graph on constant node features | The reported and implemented neighbour counts differ. The masked implementation also lacks patient-specific input variation. |

## Appendix B Dated experiment declarations

D1-D12 list the diagnostic declarations and D13 the MIMIC-IV baseline runs. The declaration files, their dates and SHA-256 hashes, and the result records are in Supplementary Data S1. Two declarations were saved shortly after launch but before reported outcomes. English translations accompany the Chinese-language declarations D8-D12. D13 is a separate declaration for the four comparison methods; it is identified separately because it is not one of the 12 diagnostic declarations counted above.

**Table B1.** Diagnostic experiment record. Early outcomes are interpreted in light of the later finding of contaminated training membership. In particular, D4-D5 near-null attack contrasts on contaminated outputs are not evidence of privacy; D5 includes one ACTG 175 detection. The capped MIMIC-IV result in D9 was followed by the full-data evaluation in D11.

| ID | Experiment | Outcome | Frozen before launch |
|---|---|---|---|
| D1 | Sampling-time perturbation and neighbour count as privacy controls (contaminated post-mask pipeline) | Declared criterion not met on the evaluated contaminated outputs; this does not establish a valid membership-risk threshold. | yes |
| D2 | Training-time perturbation grid, ACTG 175 and WBCD (contaminated post-mask pipeline) | Declared prediction not met; grid not extended. Membership interpretation is limited by later-identified contamination. | no: saved 33 s after launch |
| D3 | Training-time perturbation grid, MIMIC-IV (contaminated post-mask pipeline) | No converged setting met the declared target; one setting diverged. Training membership was subsequently found contaminated. | no: saved 77 s after a launch that failed; rerun hours later |
| D4 | Distance and density-ratio attacks on contaminated-pipeline outputs | None of 15 TabGraphSyn configurations exceeded its cohort's resolution under the distance attack; positive controls and TabDDPM (ACTG 175, TCGA) detected; density-ratio attack discarded after failing its positive control. | yes |
| D5 | MIA-EPT on contaminated-pipeline outputs | One TabGraphSyn configuration exceeded its cohort's resolution (ACTG 175, no perturbation: 0.130 against 0.110); none on TCGA, WBCD or MIMIC-IV; positive controls and TabDDPM (ACTG 175) detected. Six deviations recorded, two added after partial results. | yes |
| D6 | Probe of the conditioning channel (contaminated post-mask pipeline) | Full-table conditions and source-linked proximity exposed holdout participation. The test used evaluator access to source indices. | yes |
| D7 | Fast feasibility checks before retraining, including removal of holdout-seeded records from existing outputs | Output removal increased attack contrast; equal-sized unconditional controls changed little. Train-only correction passed six implementation checks. | yes |
| D8 | Cluster-level hints p(h), train-only pipeline, four cohorts (52 runs) | Per-record signals detected, including MIMIC-IV D = 18.7. Cluster conditions reduced additional signal within threshold; TCGA utility criterion not met (+0.0017, p = 0.31). | yes |
| D9 | Granularity ladder; contaminated post-mask vs train-only pipeline; identity controls; clipped MIMIC-IV arms; MIA-EPT | TCGA signal increased with K; random-code prediction was not supported. MIA-EPT detected TCGA and ACTG 175; capped MIMIC-IV result later diagnosed in D11. | yes |
| D10 | No-conditioning backbone vs clean TabSyn | No significant TCGA difference detected. MIMIC-IV inference limited by divergence and one converged TabSyn seed. | yes |
| D11 | Re-analysis without training: decodability, MIMIC-IV non-saturated targets, MIA-EPT diagnosis and full-data rerun | TCGA decodability association rho = 0.96; both contaminated outputs exceed TRTR on six MIMIC-IV tasks. Full-data MIA-EPT detects the GNN run at 5.2 resolution units. | yes |
| D12 | Graph used only during training, TCGA | Two TCGA mixup settings did not meet the declared utility criterion; zero-mixup control matched clean TabSyn. | yes |
| D13 | MIMIC-IV comparison baselines: CTAB-GAN+, TabDDPM, CTGAN and TabDiff, new runs with documented configurations | Fitted on the 140,821 training admissions only (CTGAN on 112,656 after its internal validation split); 156,468 rows generated per model. TabDiff was prepared in the declaration and run afterwards, so it is not covered by the freeze. Results: Figure 4 and Supplementary Data S1. | yes, for the three runs launched that day |

## Supplementary Figure

**Supplementary Figure S1.**
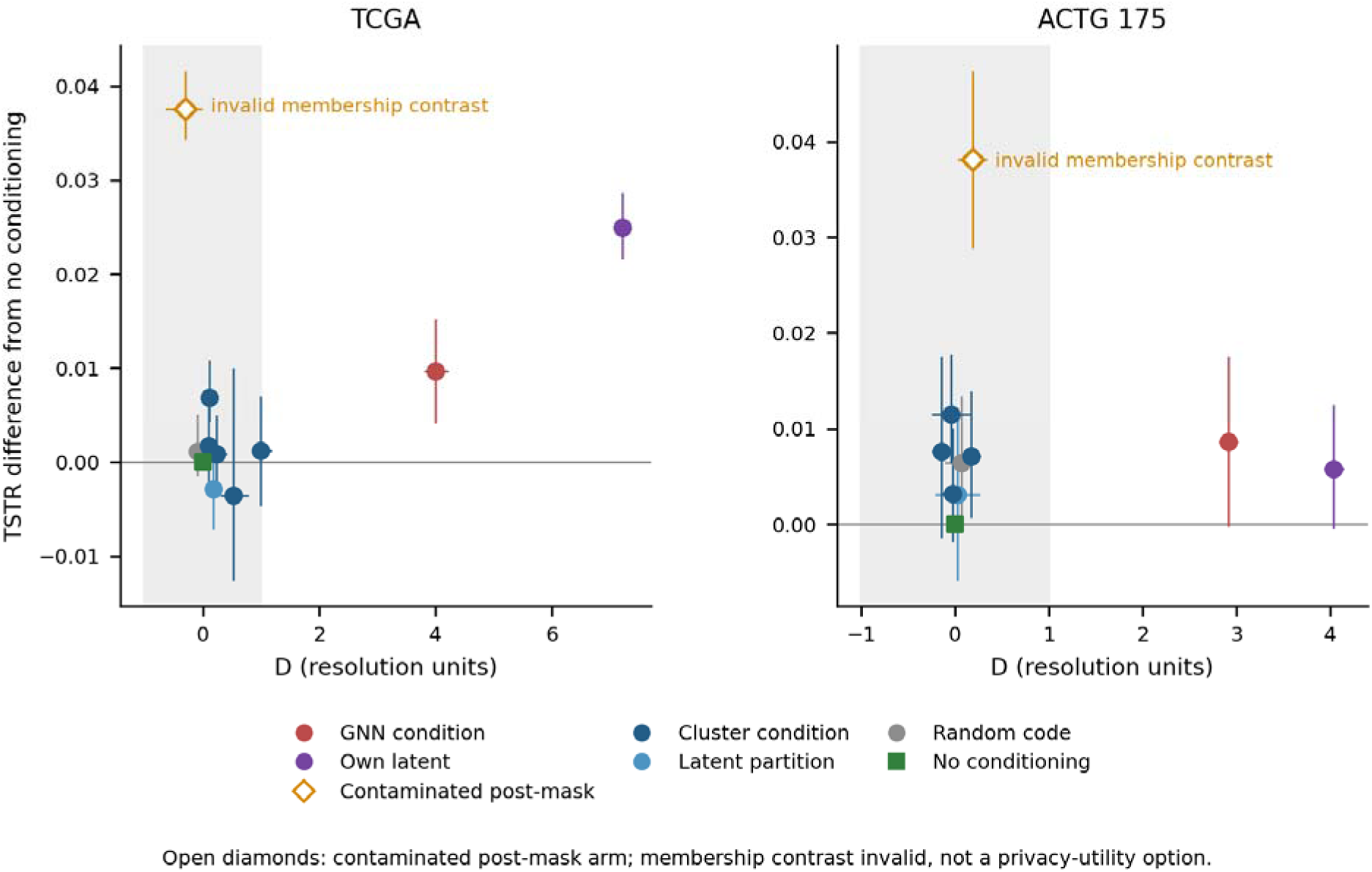
TSTR difference from the no-conditioning backbone versus additional distance-attack signal D. Points and error bars are the reported means and seed-bootstrap 95% intervals; K = 1 has one seed. TCGA has no replicated train-only arm meeting the prespecified joint criterion, while two ACTG 175 cluster arms have positive utility intervals against this backbone but no significant advantage over clean TabSyn. Contaminated-arm points are diagnostic comparisons with invalid nominal non-members, not privacy-preserving improvements.

## Supplementary Table

**Supplementary Table S1.**
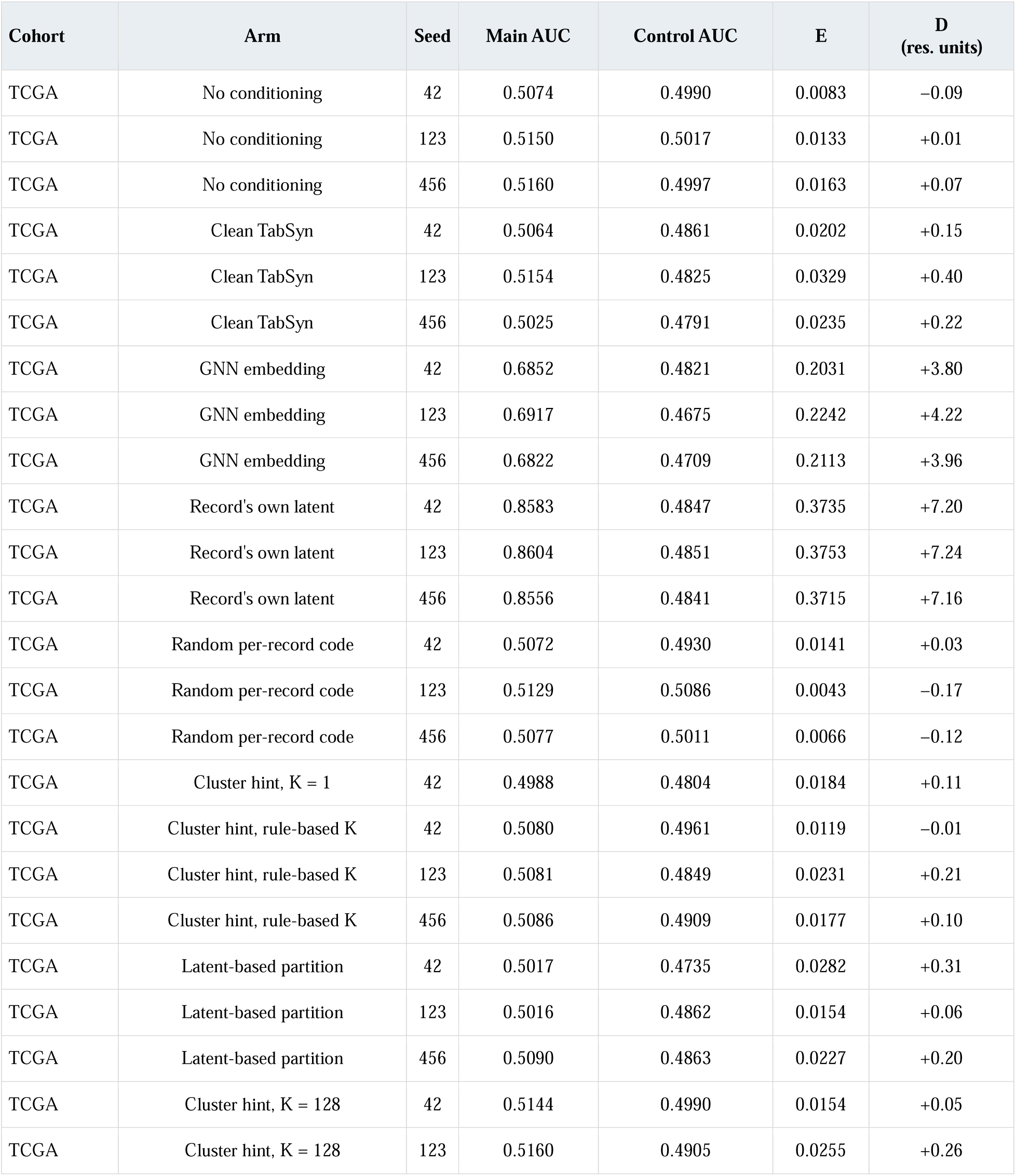
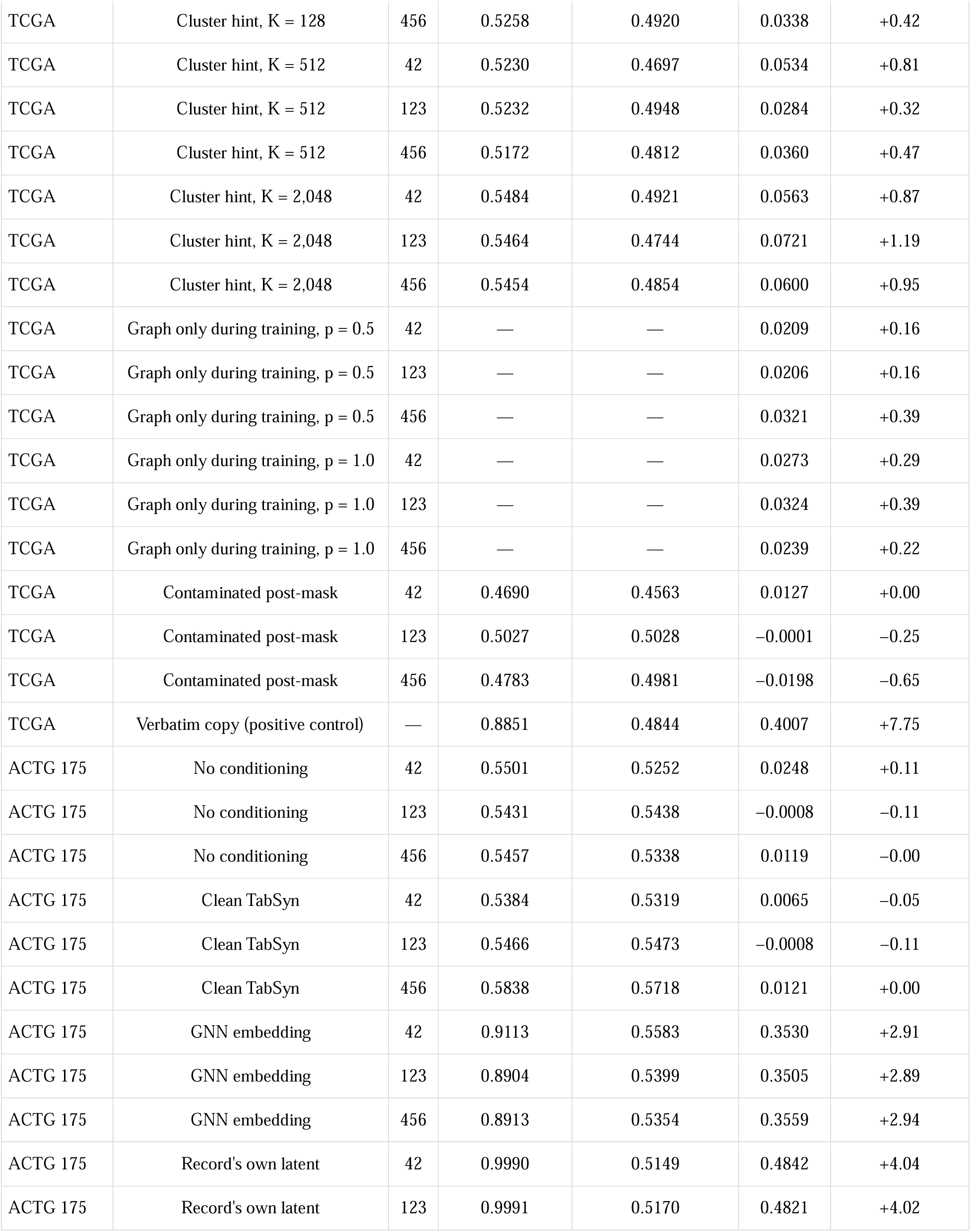
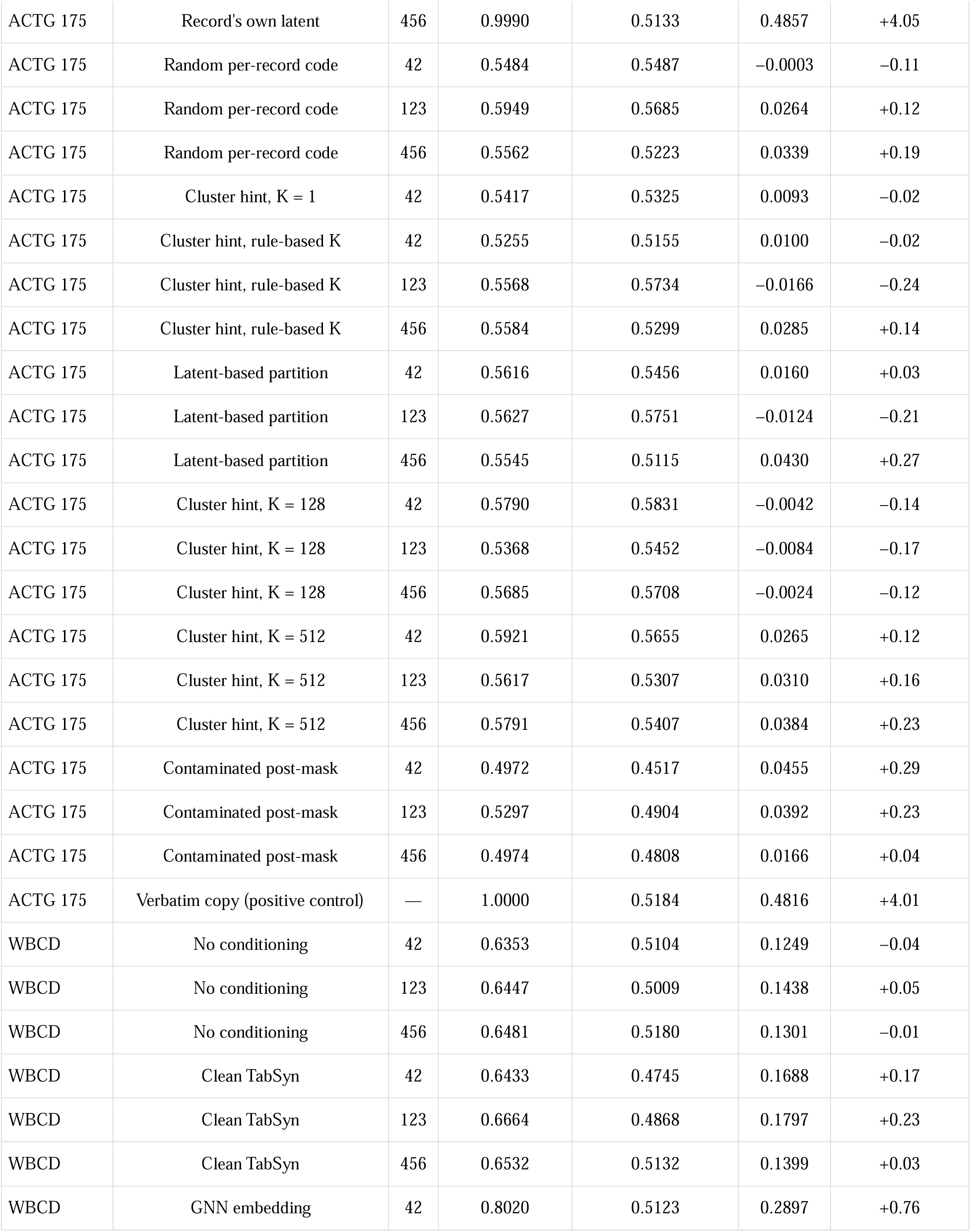
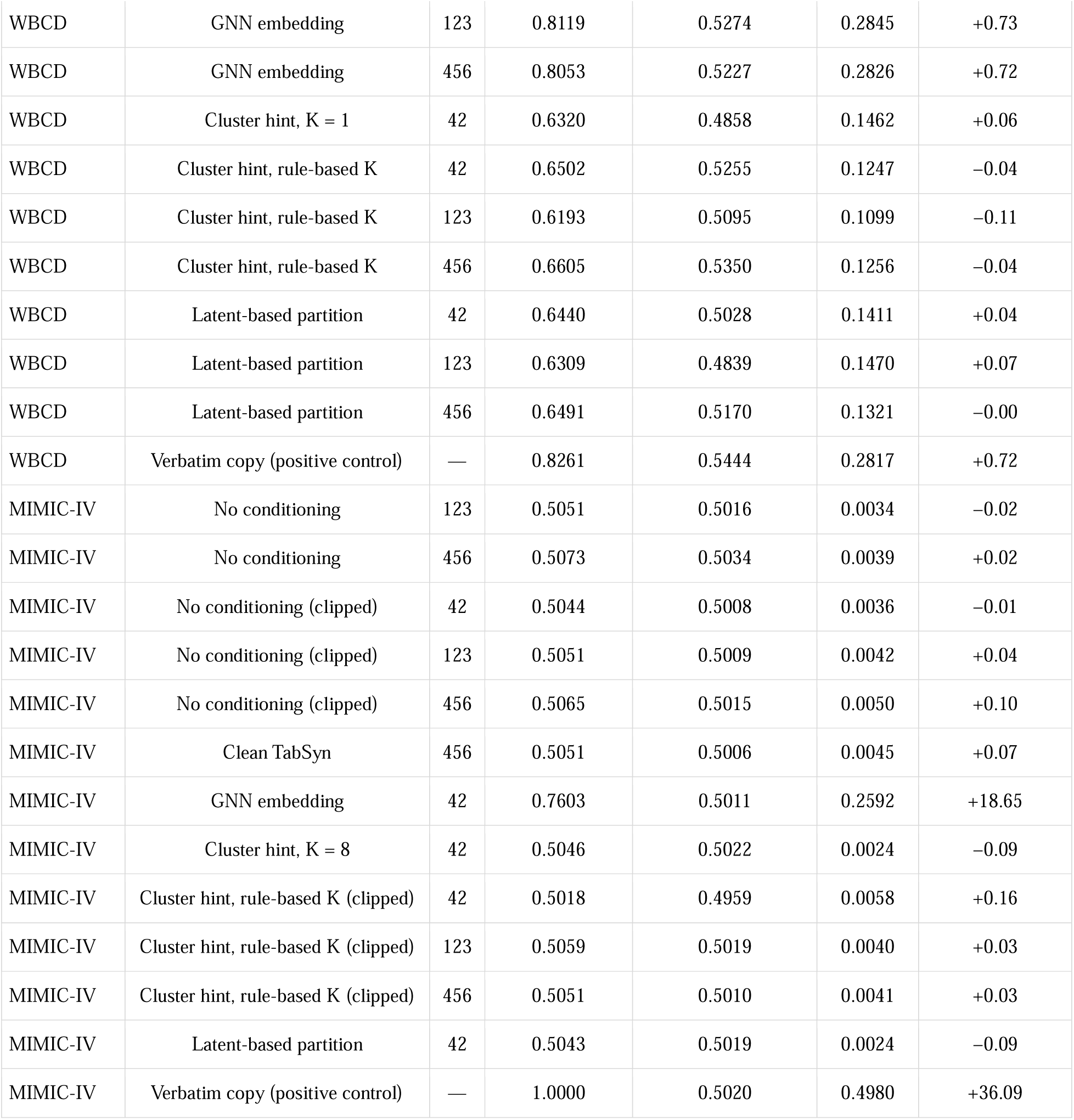
Distance attack for every analysed run. Main AUC compares training rows with H1 and control AUC compares H3 with H1; E is their difference and D = (E − mean E of the no-conditioning runs)/r, with r from Table 1. Diverged runs are excluded. For graph-only training only E was retained in the summary file, so its AUCs are not shown. WBCD values are not interpretable as membership evidence, and contaminated-arm values are nominal split contrasts (Methods).

| Cohort | Arm | Seed | Main AUC | Control AUC | E | D<br>(res. units) |
|---|---|---|---|---|---|---|
| TCGA | No conditioning | 42 | 0.5074 | 0.4990 | 0.0083 | −0.09 |
| TCGA | No conditioning | 123 | 0.5150 | 0.5017 | 0.0133 | +0.01 |
| TCGA | No conditioning | 456 | 0.5160 | 0.4997 | 0.0163 | +0.07 |
| TCGA | Clean TabSyn | 42 | 0.5064 | 0.4861 | 0.0202 | +0.15 |
| TCGA | Clean TabSyn | 123 | 0.5154 | 0.4825 | 0.0329 | +0.40 |
| TCGA | Clean TabSyn | 456 | 0.5025 | 0.4791 | 0.0235 | +0.22 |
| TCGA | GNN embedding | 42 | 0.6852 | 0.4821 | 0.2031 | +3.80 |
| TCGA | GNN embedding | 123 | 0.6917 | 0.4675 | 0.2242 | +4.22 |
| TCGA | GNN embedding | 456 | 0.6822 | 0.4709 | 0.2113 | +3.96 |
| TCGA | Record's own latent | 42 | 0.8583 | 0.4847 | 0.3735 | +7.20 |
| TCGA | Record's own latent | 123 | 0.8604 | 0.4851 | 0.3753 | +7.24 |
| TCGA | Record's own latent | 456 | 0.8556 | 0.4841 | 0.3715 | +7.16 |
| TCGA | Random per-record code | 42 | 0.5072 | 0.4930 | 0.0141 | +0.03 |
| TCGA | Random per-record code | 123 | 0.5129 | 0.5086 | 0.0043 | −0.17 |
| TCGA | Random per-record code | 456 | 0.5077 | 0.5011 | 0.0066 | −0.12 |
| TCGA | Cluster hint, K = 1 | 42 | 0.4988 | 0.4804 | 0.0184 | +0.11 |
| TCGA | Cluster hint, rule-based K | 42 | 0.5080 | 0.4961 | 0.0119 | −0.01 |
| TCGA | Cluster hint, rule-based K | 123 | 0.5081 | 0.4849 | 0.0231 | +0.21 |
| TCGA | Cluster hint, rule-based K | 456 | 0.5086 | 0.4909 | 0.0177 | +0.10 |
| TCGA | Latent-based partition | 42 | 0.5017 | 0.4735 | 0.0282 | +0.31 |
| TCGA | Latent-based partition | 123 | 0.5016 | 0.4862 | 0.0154 | +0.06 |
| TCGA | Latent-based partition | 456 | 0.5090 | 0.4863 | 0.0227 | +0.20 |
| TCGA | Cluster hint, K = 128 | 42 | 0.5144 | 0.4990 | 0.0154 | +0.05 |
| TCGA | Cluster hint, K = 128 | 123 | 0.5160 | 0.4905 | 0.0255 | +0.26 |
| TCGA | Cluster hint, K = 128 | 456 | 0.5258 | 0.4920 | 0.0338 | +0.42 |
| TCGA | Cluster hint, K = 512 | 42 | 0.5230 | 0.4697 | 0.0534 | +0.81 |
| TCGA | Cluster hint, K = 512 | 123 | 0.5232 | 0.4948 | 0.0284 | +0.32 |
| TCGA | Cluster hint, K = 512 | 456 | 0.5172 | 0.4812 | 0.0360 | +0.47 |
| TCGA | Cluster hint, K = 2,048 | 42 | 0.5484 | 0.4921 | 0.0563 | +0.87 |
| TCGA | Cluster hint, K = 2,048 | 123 | 0.5464 | 0.4744 | 0.0721 | +1.19 |
| TCGA | Cluster hint, K = 2,048 | 456 | 0.5454 | 0.4854 | 0.0600 | +0.95 |
| TCGA | Graph only during training, p = 0.5 | 42 | — | — | 0.0209 | +0.16 |
| TCGA | Graph only during training, p = 0.5 | 123 | — | — | 0.0206 | +0.16 |
| TCGA | Graph only during training, p = 0.5 | 456 | — | — | 0.0321 | +0.39 |
| TCGA | Graph only during training, p = 1.0 | 42 | — | — | 0.0273 | +0.29 |
| TCGA | Graph only during training, p = 1.0 | 123 | — | — | 0.0324 | +0.39 |
| TCGA | Graph only during training, p = 1.0 | 456 | — | — | 0.0239 | +0.22 |
| TCGA | Contaminated post-mask | 42 | 0.4690 | 0.4563 | 0.0127 | +0.00 |
| TCGA | Contaminated post-mask | 123 | 0.5027 | 0.5028 | −0.0001 | −0.25 |
| TCGA | Contaminated post-mask | 456 | 0.4783 | 0.4981 | −0.0198 | −0.65 |
| TCGA | Verbatim copy (positive control) | — | 0.8851 | 0.4844 | 0.4007 | +7.75 |
| ACTG 175 | No conditioning | 42 | 0.5501 | 0.5252 | 0.0248 | +0.11 |
| ACTG 175 | No conditioning | 123 | 0.5431 | 0.5438 | −0.0008 | −0.11 |
| ACTG 175 | No conditioning | 456 | 0.5457 | 0.5338 | 0.0119 | −0.00 |
| ACTG 175 | Clean TabSyn | 42 | 0.5384 | 0.5319 | 0.0065 | −0.05 |
| ACTG 175 | Clean TabSyn | 123 | 0.5466 | 0.5473 | −0.0008 | −0.11 |
| ACTG 175 | Clean TabSyn | 456 | 0.5838 | 0.5718 | 0.0121 | +0.00 |
| ACTG 175 | GNN embedding | 42 | 0.9113 | 0.5583 | 0.3530 | +2.91 |
| ACTG 175 | GNN embedding | 123 | 0.8904 | 0.5399 | 0.3505 | +2.89 |
| ACTG 175 | GNN embedding | 456 | 0.8913 | 0.5354 | 0.3559 | +2.94 |
| ACTG 175 | Record's own latent | 42 | 0.9990 | 0.5149 | 0.4842 | +4.04 |
| ACTG 175 | Record's own latent | 123 | 0.9991 | 0.5170 | 0.4821 | +4.02 |
| ACTG 175 | Record's own latent | 456 | 0.9990 | 0.5133 | 0.4857 | +4.05 |
| ACTG 175 | Random per-record code | 42 | 0.5484 | 0.5487 | −0.0003 | −0.11 |
| ACTG 175 | Random per-record code | 123 | 0.5949 | 0.5685 | 0.0264 | +0.12 |
| ACTG 175 | Random per-record code | 456 | 0.5562 | 0.5223 | 0.0339 | +0.19 |
| ACTG 175 | Cluster hint, K = 1 | 42 | 0.5417 | 0.5325 | 0.0093 | −0.02 |
| ACTG 175 | Cluster hint, rule-based K | 42 | 0.5255 | 0.5155 | 0.0100 | −0.02 |
| ACTG 175 | Cluster hint, rule-based K | 123 | 0.5568 | 0.5734 | −0.0166 | −0.24 |
| ACTG 175 | Cluster hint, rule-based K | 456 | 0.5584 | 0.5299 | 0.0285 | +0.14 |
| ACTG 175 | Latent-based partition | 42 | 0.5616 | 0.5456 | 0.0160 | +0.03 |
| ACTG 175 | Latent-based partition | 123 | 0.5627 | 0.5751 | −0.0124 | −0.21 |
| ACTG 175 | Latent-based partition | 456 | 0.5545 | 0.5115 | 0.0430 | +0.27 |
| ACTG 175 | Cluster hint, K = 128 | 42 | 0.5790 | 0.5831 | −0.0042 | −0.14 |
| ACTG 175 | Cluster hint, K = 128 | 123 | 0.5368 | 0.5452 | −0.0084 | −0.17 |
| ACTG 175 | Cluster hint, K = 128 | 456 | 0.5685 | 0.5708 | −0.0024 | −0.12 |
| ACTG 175 | Cluster hint, K = 512 | 42 | 0.5921 | 0.5655 | 0.0265 | +0.12 |
| ACTG 175 | Cluster hint, K = 512 | 123 | 0.5617 | 0.5307 | 0.0310 | +0.16 |
| ACTG 175 | Cluster hint, K = 512 | 456 | 0.5791 | 0.5407 | 0.0384 | +0.23 |
| ACTG 175 | Contaminated post-mask | 42 | 0.4972 | 0.4517 | 0.0455 | +0.29 |
| ACTG 175 | Contaminated post-mask | 123 | 0.5297 | 0.4904 | 0.0392 | +0.23 |
| ACTG 175 | Contaminated post-mask | 456 | 0.4974 | 0.4808 | 0.0166 | +0.04 |
| ACTG 175 | Verbatim copy (positive control) | — | 1.0000 | 0.5184 | 0.4816 | +4.01 |
| WBCD | No conditioning | 42 | 0.6353 | 0.5104 | 0.1249 | −0.04 |
| WBCD | No conditioning | 123 | 0.6447 | 0.5009 | 0.1438 | +0.05 |
| WBCD | No conditioning | 456 | 0.6481 | 0.5180 | 0.1301 | −0.01 |
| WBCD | Clean TabSyn | 42 | 0.6433 | 0.4745 | 0.1688 | +0.17 |
| WBCD | Clean TabSyn | 123 | 0.6664 | 0.4868 | 0.1797 | +0.23 |
| WBCD | Clean TabSyn | 456 | 0.6532 | 0.5132 | 0.1399 | +0.03 |
| WBCD | GNN embedding | 42 | 0.8020 | 0.5123 | 0.2897 | +0.76 |
| WBCD | GNN embedding | 123 | 0.8119 | 0.5274 | 0.2845 | +0.73 |
| WBCD | GNN embedding | 456 | 0.8053 | 0.5227 | 0.2826 | +0.72 |
| WBCD | Cluster hint, K = 1 | 42 | 0.6320 | 0.4858 | 0.1462 | +0.06 |
| WBCD | Cluster hint, rule-based K | 42 | 0.6502 | 0.5255 | 0.1247 | −0.04 |
| WBCD | Cluster hint, rule-based K | 123 | 0.6193 | 0.5095 | 0.1099 | −0.11 |
| WBCD | Cluster hint, rule-based K | 456 | 0.6605 | 0.5350 | 0.1256 | −0.04 |
| WBCD | Latent-based partition | 42 | 0.6440 | 0.5028 | 0.1411 | +0.04 |
| WBCD | Latent-based partition | 123 | 0.6309 | 0.4839 | 0.1470 | +0.07 |
| WBCD | Latent-based partition | 456 | 0.6491 | 0.5170 | 0.1321 | −0.00 |
| WBCD | Verbatim copy (positive control) | — | 0.8261 | 0.5444 | 0.2817 | +0.72 |
| MIMIC-IV | No conditioning | 123 | 0.5051 | 0.5016 | 0.0034 | −0.02 |
| MIMIC-IV | No conditioning | 456 | 0.5073 | 0.5034 | 0.0039 | +0.02 |
| MIMIC-IV | No conditioning (clipped) | 42 | 0.5044 | 0.5008 | 0.0036 | −0.01 |
| MIMIC-IV | No conditioning (clipped) | 123 | 0.5051 | 0.5009 | 0.0042 | +0.04 |
| MIMIC-IV | No conditioning (clipped) | 456 | 0.5065 | 0.5015 | 0.0050 | +0.10 |
| MIMIC-IV | Clean TabSyn | 456 | 0.5051 | 0.5006 | 0.0045 | +0.07 |
| MIMIC-IV | GNN embedding | 42 | 0.7603 | 0.5011 | 0.2592 | +18.65 |
| MIMIC-IV | Cluster hint, K = 8 | 42 | 0.5046 | 0.5022 | 0.0024 | −0.09 |
| MIMIC-IV | Cluster hint, rule-based K (clipped) | 42 | 0.5018 | 0.4959 | 0.0058 | +0.16 |
| MIMIC-IV | Cluster hint, rule-based K (clipped) | 123 | 0.5059 | 0.5019 | 0.0040 | +0.03 |
| MIMIC-IV | Cluster hint, rule-based K (clipped) | 456 | 0.5051 | 0.5010 | 0.0041 | +0.03 |
| MIMIC-IV | Latent-based partition | 42 | 0.5043 | 0.5019 | 0.0024 | −0.09 |
| MIMIC-IV | Verbatim copy (positive control) | — | 1.0000 | 0.5020 | 0.4980 | +36.09 |

