## Supplementary figures and images for "Revisiting TabGraphSyn: training boundaries and record-level conditioning in synthetic clinical data evaluation"

### Figure1_framework.png

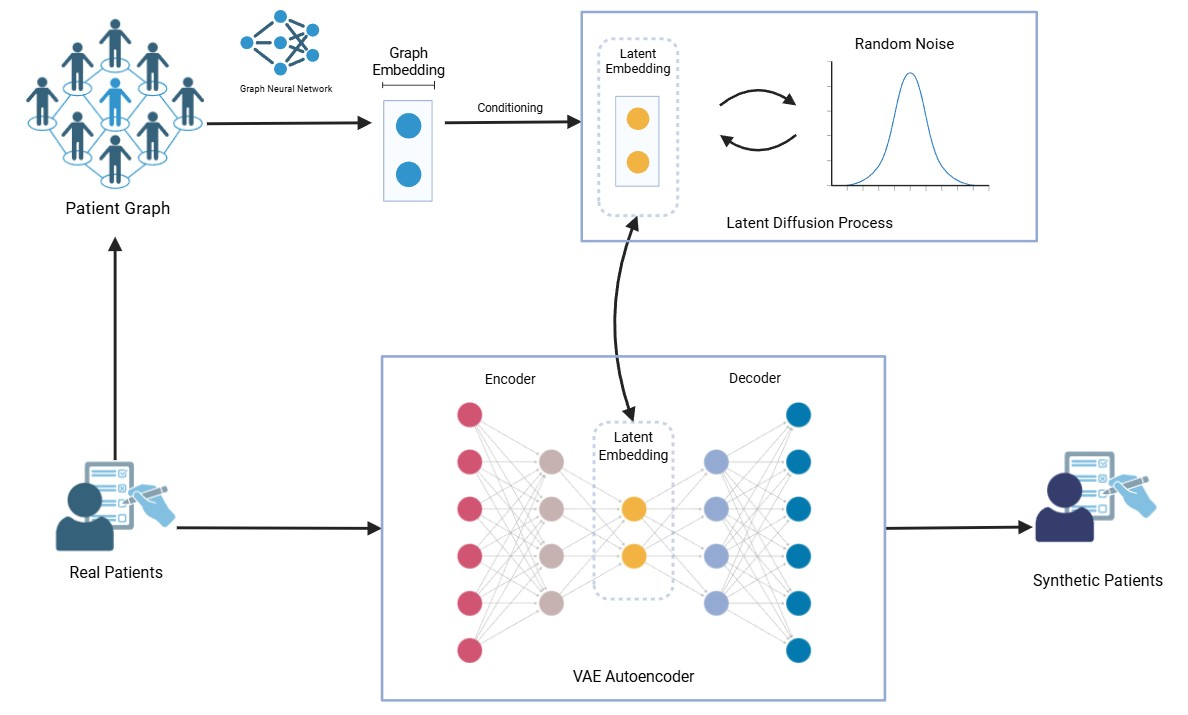

### Figure2_output_removal.png

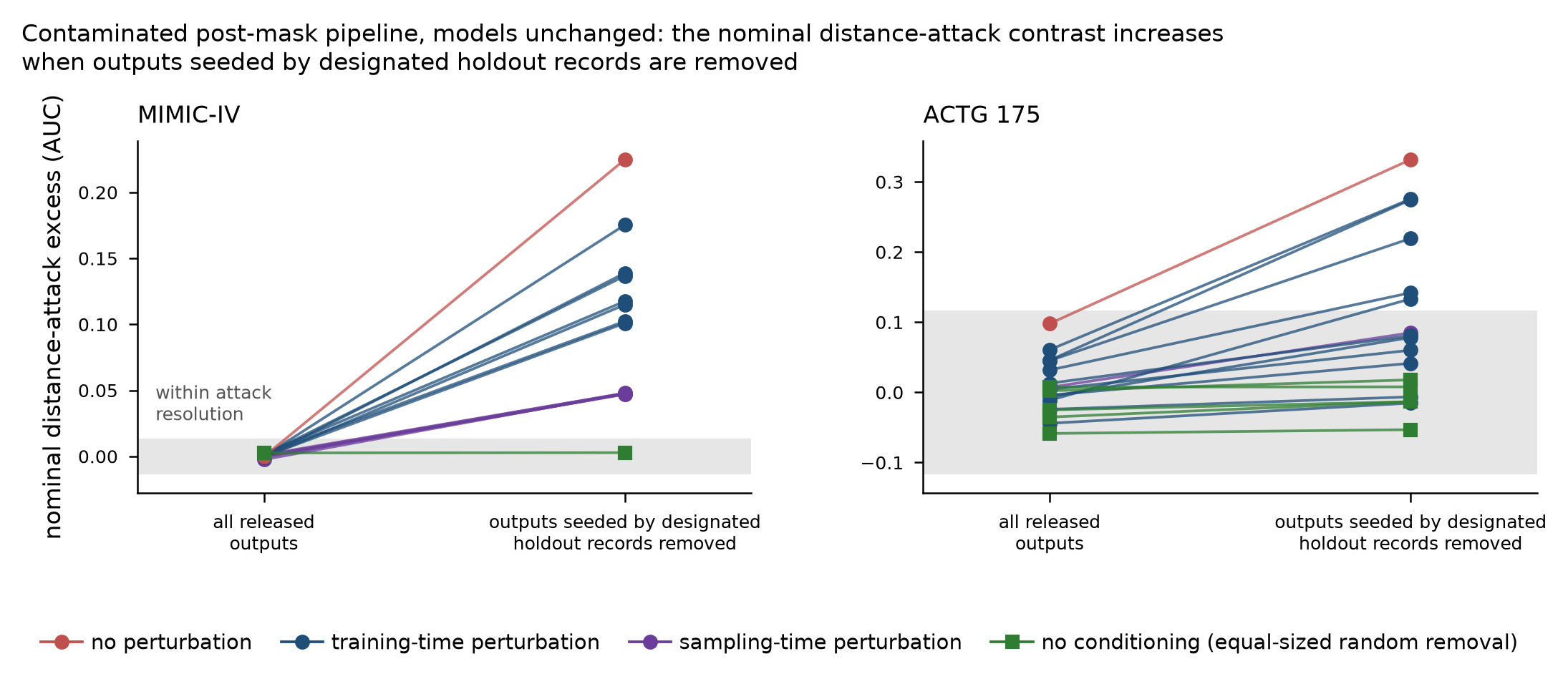

### Figure3_paired.png

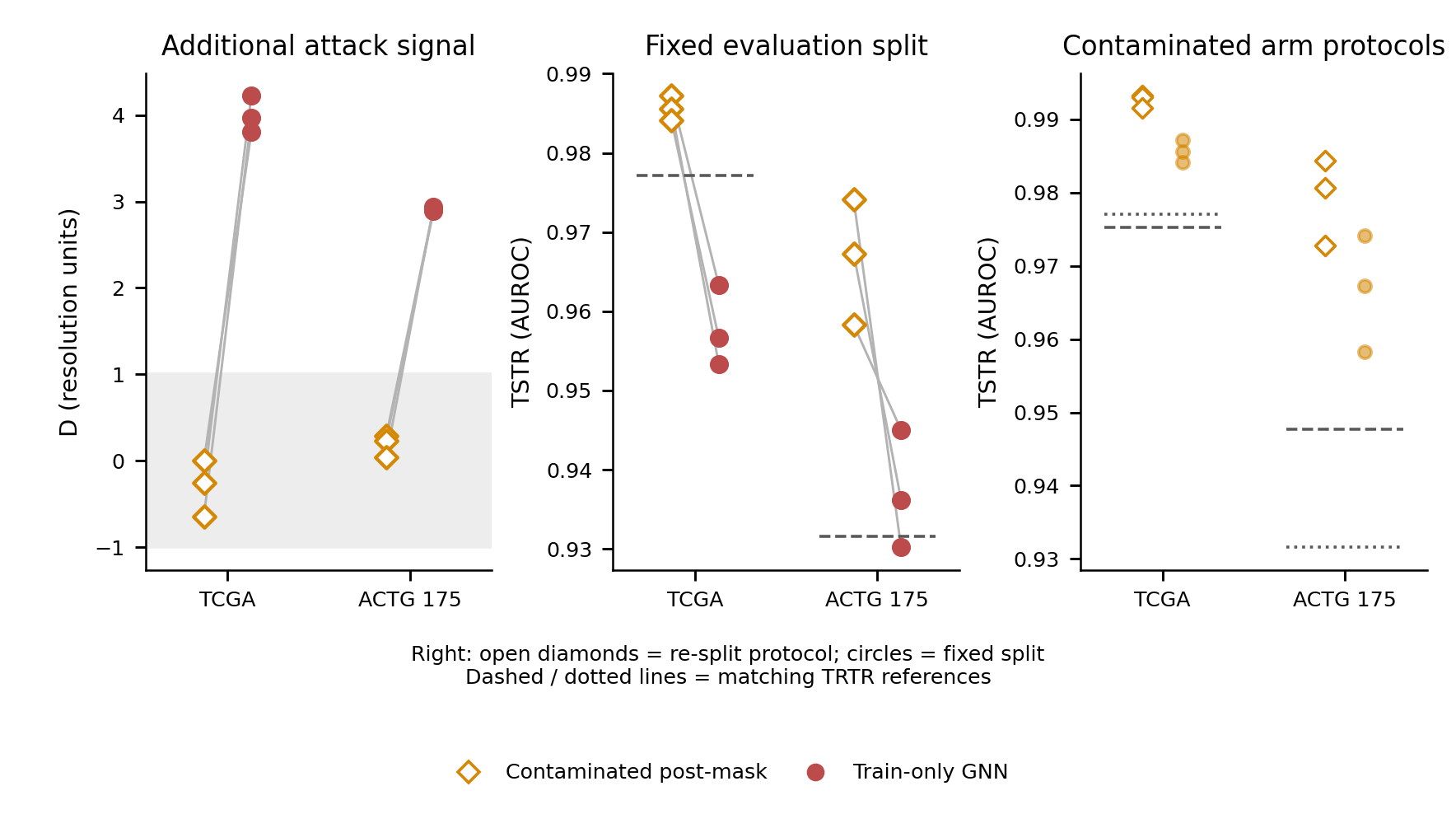

### Figure4_mimic_targets.png

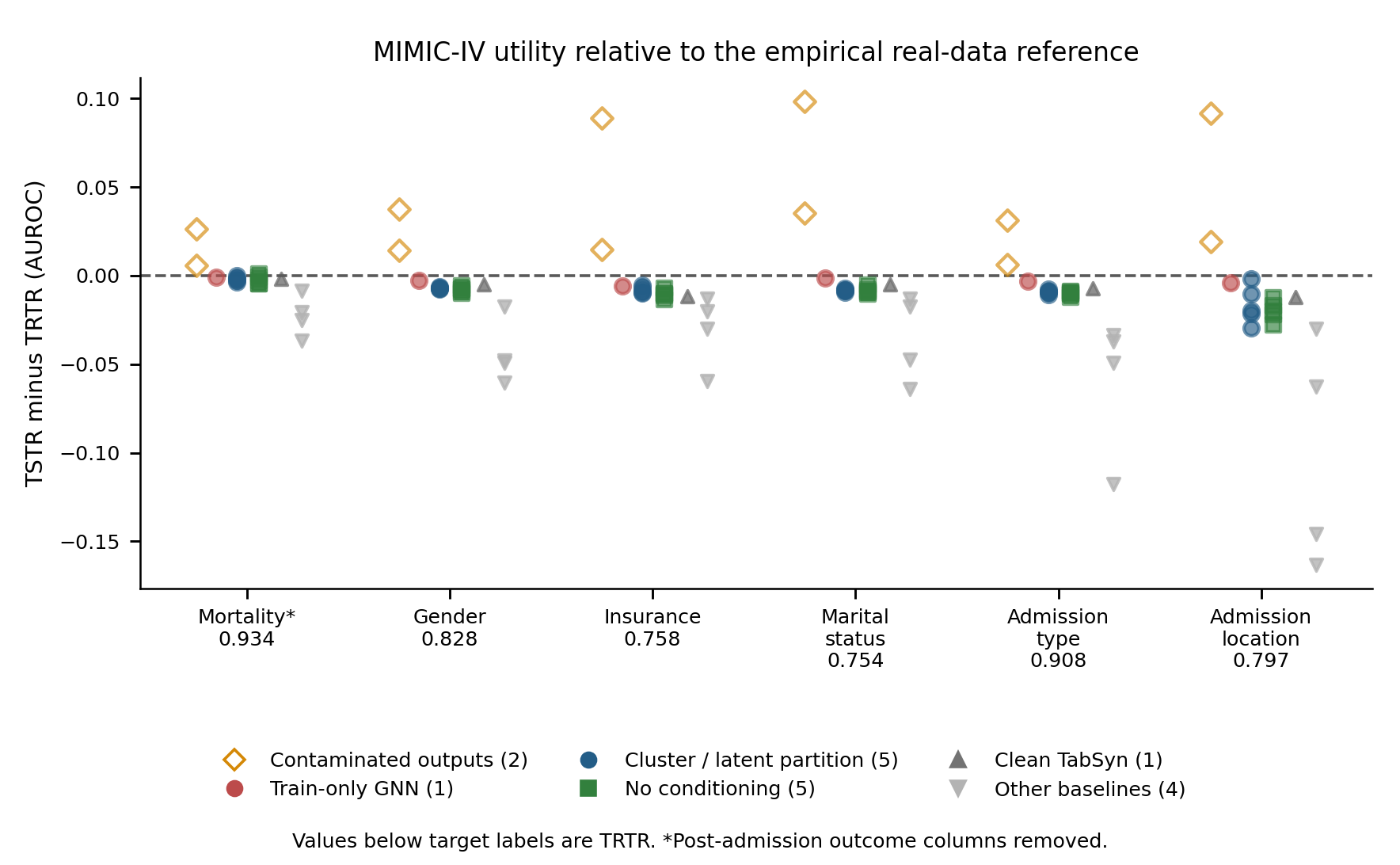

### Figure5_decodability.png

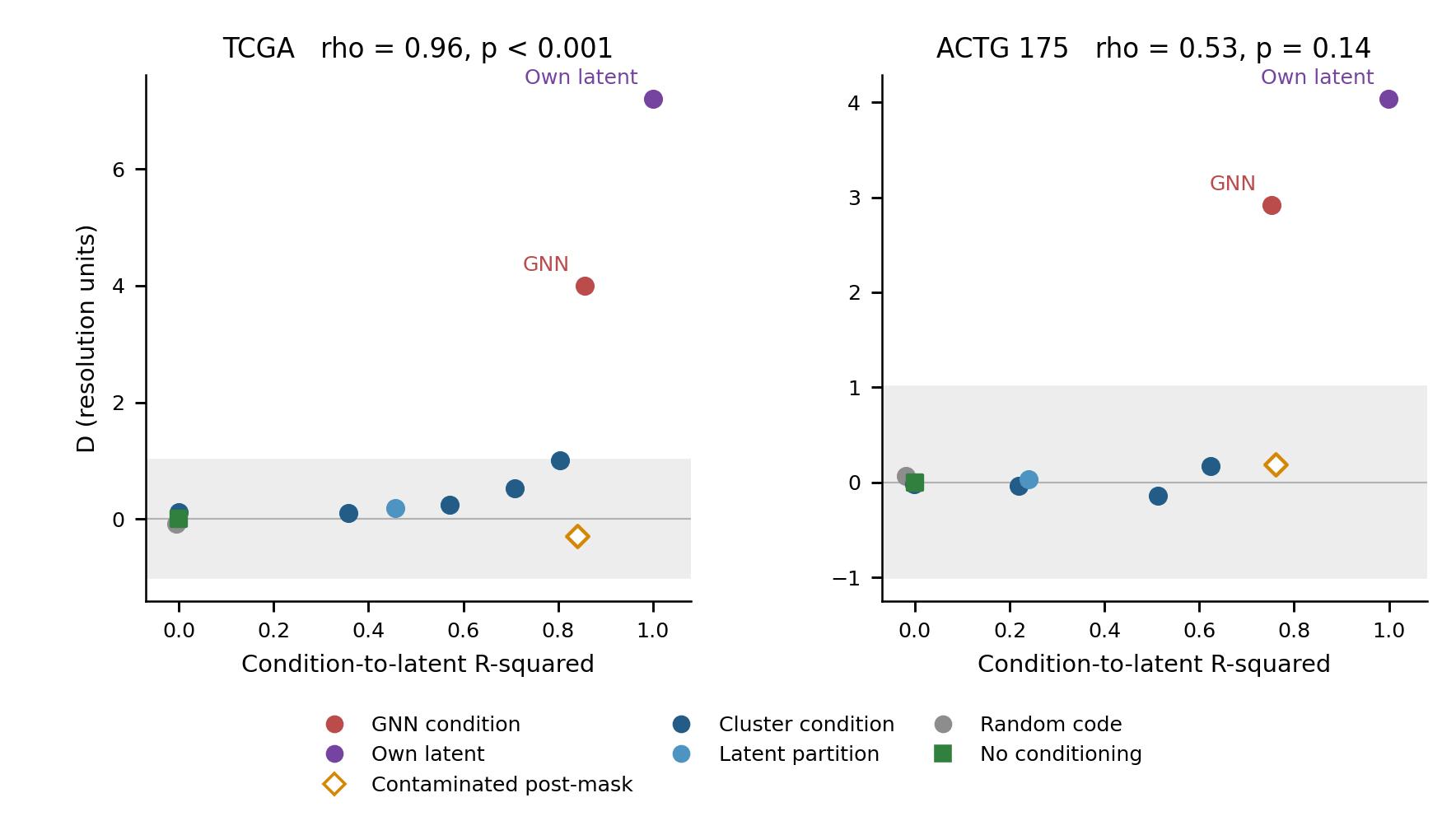

### Figure6_granularity.png

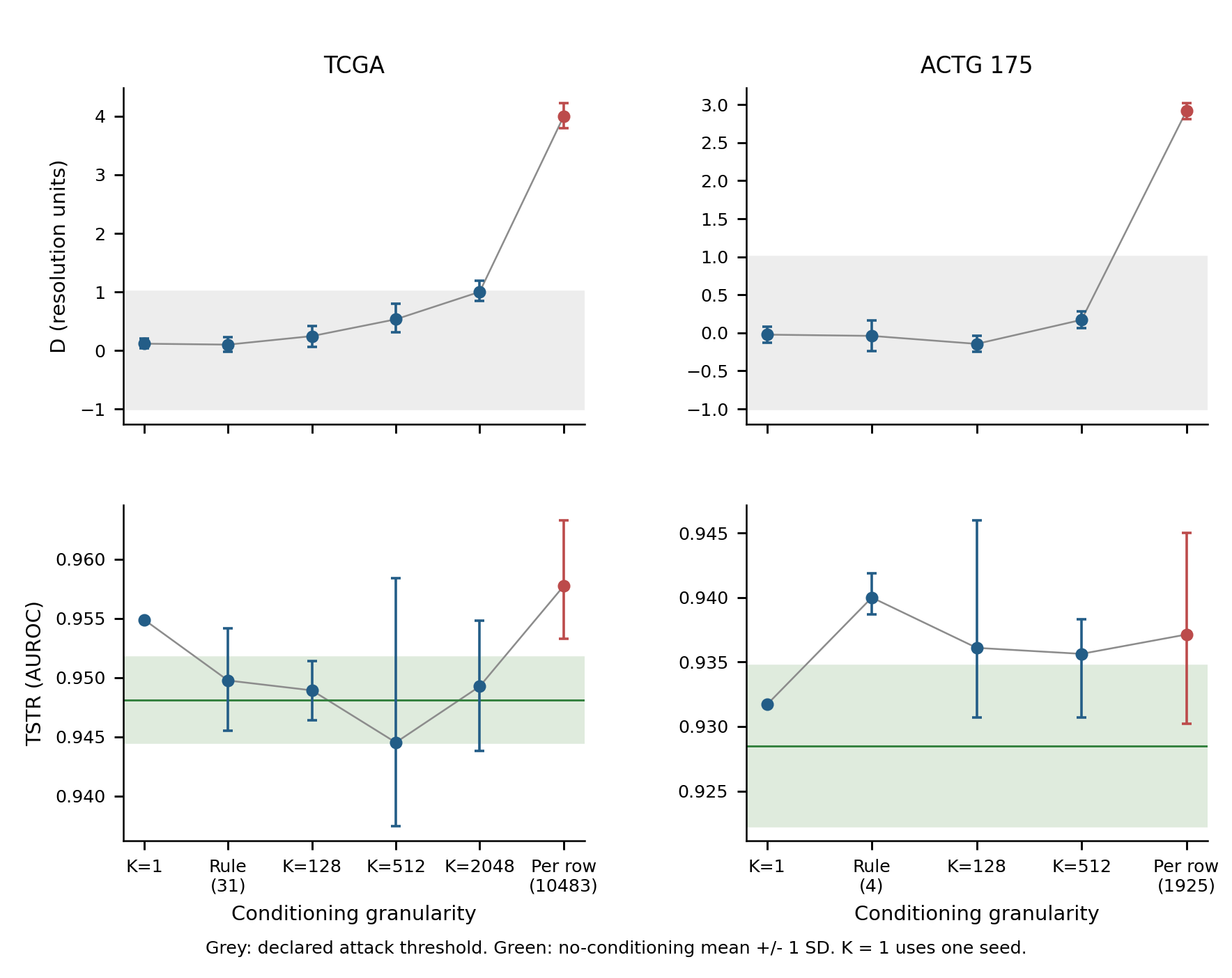

### Figure7_vs_clean_tabsyn.png

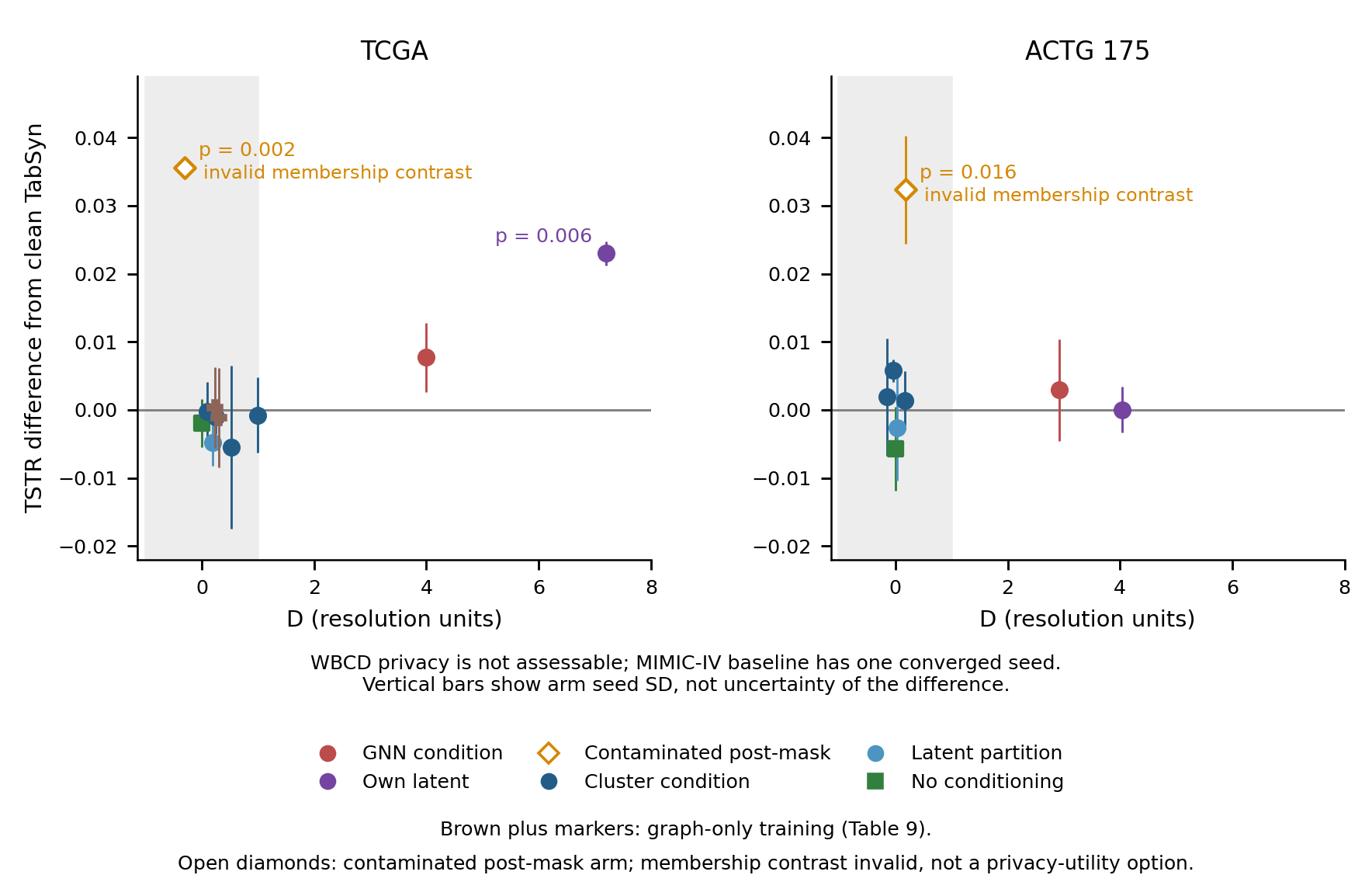

### FigureS1_vs_no_conditioning.png

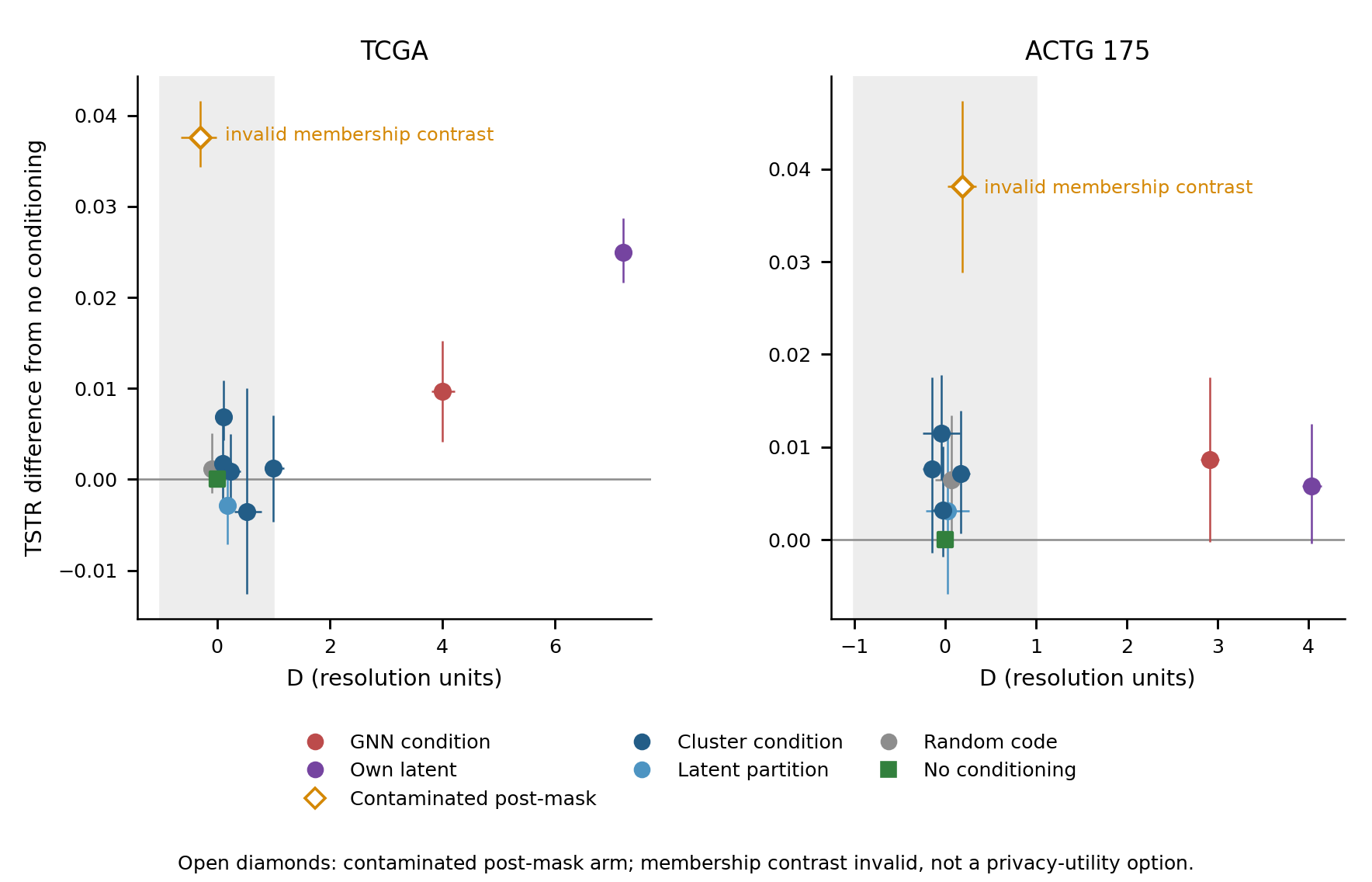
